# Transcriptional pathways of definitive ALS genes implicate novel disease-associated target genes

**DOI:** 10.64898/2026.07.21.26358605

**Authors:** Michael R. Fiorini, Allison A. Dilliott, Sami Alsabagh, Nadege Wredenhagen, Sali M.K. Farhan

**Affiliations:** Department of Human Genetics, McGill University, Montreal, Quebec H3A 0C7, Canada; The Montreal Neurological Institute-Hospital, McGill University, Montreal, Quebec H3A 2B4, Canada; Faculty of Science and Schulich School of Medicine & Dentistry, Western University, London, Ontario, N6A 5C1; Department of Neurology and Neurosurgery, McGill University, Montreal, Quebec H3A 1Y2, Canada

**Keywords:** Amyotrophic lateral sclerosis, induced pluripotent stem cell, RNAseq, co-expression network analysis, rare variant gene burden

## Abstract

Amyotrophic lateral sclerosis (ALS) is a progressive neurodegenerative disease characterized by the selective degeneration of motor neurons with a substantial genetic contribution to disease risk. Distinct genetic etiologies converge on key pathogenic processes in ALS, including aberrant RNA metabolism, proteostatic stress, dysregulated vesicular transport, and mitochondrial dysfunction. Here, we investigate whether perturbed transcriptional programs associated with definitive ALS genes harbor elusive genetic contributors to disease.

We analyzed RNA sequencing data profiling induced pluripotent stem cell-derived motor neurons from individuals with ALS (*N* = 438) and controls (*N* = 187). Leveraging an integrative analytical framework, we identified transcriptional pathways linking variants in definitive ALS genes to widespread coordinated co-expression activity driven by gene-specific transcriptional programs. We subsequently subjected all genes comprising significant pathways to genetic analyses to uncover novel targets and characterized these genes using single-nuclei transcriptomics of the motor and prefrontal cortices of individuals with ALS and controls.

The identified transcriptional programs associated with definitive ALS genes showed strong concordance to established biology, including dysregulated proteostasis and mitochondrial stress in *SOD1* ALS, protein aggregation and degradation pathways in *FUS* ALS, and altered vesicle dynamics, synaptic transmission, autophagy, and trafficking programs in *GRN* ALS. These pathways encompassed both well-established ALS genes, as well as six target genes—*PTPRN2*, *UNC13C*, *TTC3*, *USP10*, *PSMD4*, and *RUFY3*—of which *USP10*, *PSMD4,* and *RUFY3* have not yet been described in the context of ALS genetic association studies. Single-nuclei analyses demonstrated that target gene expression was enriched in the baseline architecture of neuronal populations, supporting neuron-intrinsic mechanisms of vulnerability. *PSMD4* showed region-specific dysregulation in selectively vulnerable Layer 5 motor neurons of individuals with ALS, implicating regionally divergent proteasome activation as a neuron-intrinsic feature of ALS vulnerability.

Collectively, this work advances the molecular understanding of distinct genetic ALS etiologies, provides mechanistic insight into emerging ALS target genes, and establishes a generalizable framework that is readily applicable across heritable diseases for the identification of disease-relevant molecular pathways and target genes.

## Introduction

Amyotrophic lateral sclerosis (ALS) is a fatal neurodegenerative disease characterized by progressive degeneration of upper and lower motor neurons^1^. A clear familial history is observed in ∼10% of individuals with ALS, whereas the remaining ∼90% of cases are considered genetically complex, with disease heritability estimates of ∼40-60%^2,3^. Over 50 genes have been implicated in ALS through genetic studies^4^. Of these, approximately half are now considered to have a definitive association with the disease, including *SOD1*, *FUS*, *NEK1*, *TBK1*, *OPTN*, and pathogenic repeat expansions in *C9orf72* and *ATXN2*^4^. Despite considerable progress, known genetic factors explain only ∼5-10% of disease heritability^5^, highlighting the likelihood of additional, undiscovered genetic contributors.

Defining the genetic architecture of ALS is essential for accurate diagnosis, mechanistic insight, and therapeutic development, as exemplified by the approval of tofersen for *SOD1*-mutation-positive ALS^6^. Substantial evidence indicates that a core set of interconnected biological processes are disrupted in ALS, driven by mutations in established disease genes, including aberrant RNA metabolism and nucleocytoplasmic transport (*ATXN2*, *C9orf72, FUS*, *TARDBP*), protein aggregation and impaired proteostasis (*ALS2*, *C9orf72*, *FUS*, *OPTN*, *SOD1*, *TBK1*, *UBQLN2*, *VAPB*, *VCP*), vesicle trafficking and lysosomal dysfunction (*C9orf72*, *GRN*), oxidative stress (*ALS2*, *SOD1*, *TARDBP*), and mitochondrial dysfunction (*CHCHD10*, *SOD1*, *TARDBP*)^3,7^. The convergence of key pathogenic processes across genetic etiologies raises the possibility that these pathways harbor novel genetic factors influencing disease risk.

Here, we investigated whether transcriptional pathways associated with definitive ALS genes encompass novel genetic risk factors for the disease. Leveraging bulk RNA sequencing (RNAseq) of induced pluripotent stem cell-(iPSC) derived motor neurons from individuals with ALS and controls, we applied an integrative analytical framework to model transcriptional perturbations across genetic ALS subtypes (**Fig. 1**). We provided a systems-level characterization of the molecular consequences of variants in ALS genes and identified novel disease-associated target genes embedded within the transcriptional pathways. Finally, we analyzed an orthogonal single-nuclei RNA sequencing (snRNAseq) dataset to provide mechanistic insight into the potential role of novel target genes in disease pathogenesis.

**Fig. 1.**
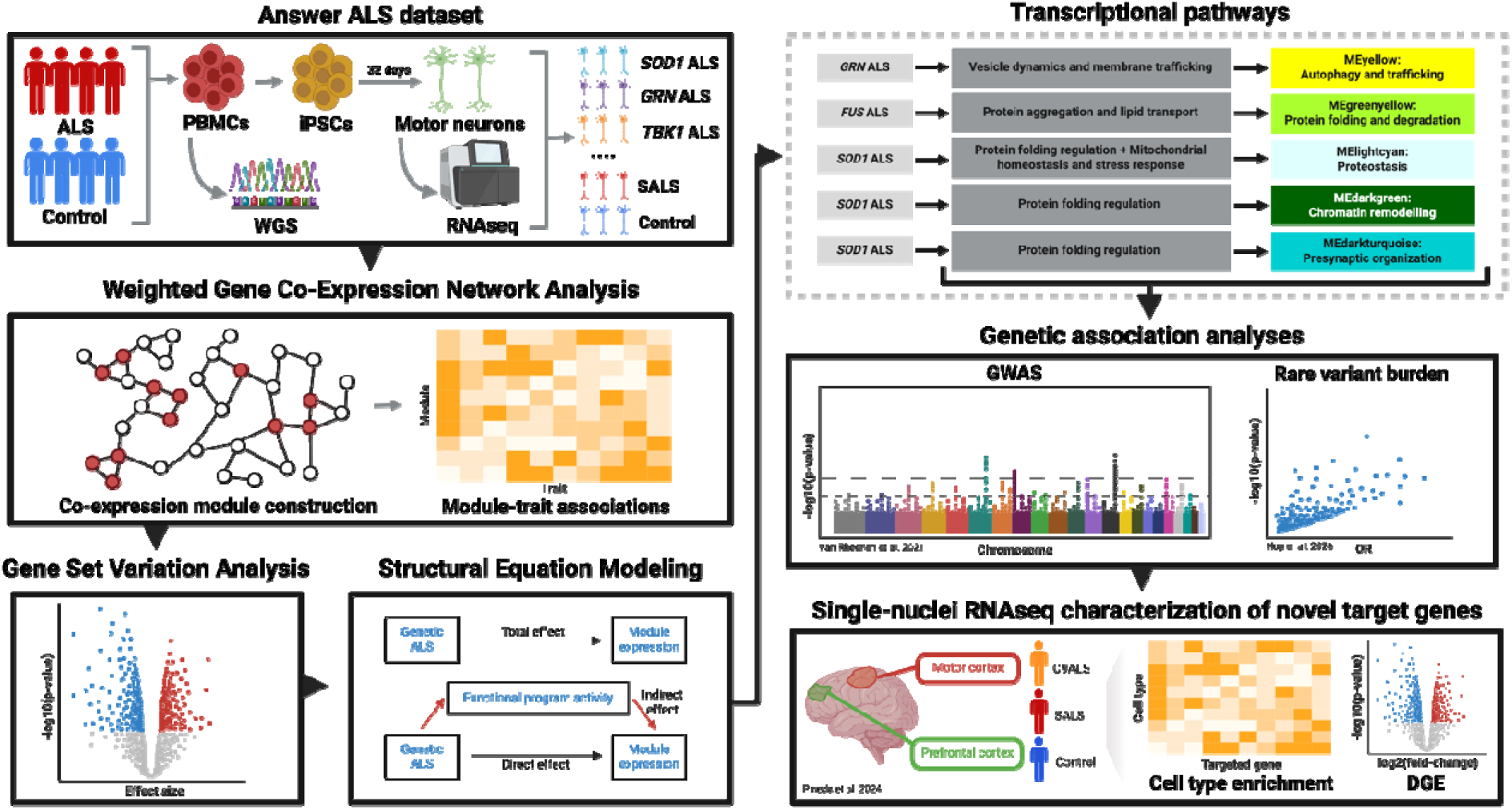
Schematic overview of the analytical workflow. ALS samples from the Answer ALS dataset (*N* = 438) were first stratified by carrier status of variants in known ALS-associated genes. We then performed weighted gene co-expression network analysis to identify co-expression modules associated with variant carrier groups. To nominate biological processe underlying aberrant module activity, we applied module-independent gene set variation analysis, followed by mediation analysis. This approach identified statistically significant transcriptional pathways linking variants in ALS genes to aberrant co-expression module activity. Genes within the significantly associated pathways were systematically evaluated using data from existing large-scale common and rare variant ALS genetic association studies to nominate novel target disease genes. Target genes were characterized using a single-nuclei RNA sequencing dataset profiling the motor and prefrontal cortices of individuals with *C9orf72* ALS (C9ALS; *N* = 16), sporadic ALS (SALS; *N* = 17), and controls (*N* = 16).

## Materials and Methods

### Sample stratification based on variant carrier status in ALS genes

Bulk RNAseq of iPSC-derived motor neurons and whole-genome sequencing (WGS) of peripheral blood mononuclear cells (PBMC) from 438 individuals with ALS and 187 controls were obtained from the Answer ALS data portal (January 2025; https://dataportal.answerals.org/)^8^. Genetic data were processed using *Segpy* (v1.0.0) to identify variant carriers^9^. Variants were annotated with the Ensembl Variant Effect Predictor (VEP; January 2025) to obtain allele frequency estimates from the gnomAD exome and genome reference populations^10,11^. Individuals with ALS were classified as variant carriers if they: (i) carried a missense variant (missense substitutions or in-frame insertions/deletions) or putative loss-of-function (pLoF) variant (stop-gain/loss, frameshift insertions/deletions, or splice-region variants—defined as variants within splice regions with a SpliceAI score > 0.2) with a minor allele frequency (MAF) <0.1% in genes classified as moderate, strong, or definitive by the ClinGen ALS Gene Curation Expert Panel (GCEP), or (ii) carried a missense variant with a REVEL score > 0.644 or pLoF and MAF < 0.1% in genes classified as limited by the ALS GCEP^12–14^. For *SPG11* and *ALS2*, only individuals with homozygous variants were classified as carriers due to the established autosomal recessive inheritance of these genes^15,16^. Genome data were analyzed using *ExpansionHunter* to identify known disease-associated trinucleotide repeats in *ATXN2* and *C9orf72*^17^. Individuals with ALS with repeat expansions >30 in *C9orf72* and ≥30 in *ATXN2* were also classified as carriers^18^. Downstream analyses focused on ALS GCEP-defined definitive genes (definitive ALS genes hereafter) with >3 variant carriers in individuals with ALS.

### Weighted gene co-expression network analysis

To investigate coordinated transcriptional programs across variant carrier groups, we performed weighted gene co-expression network analysis (WGCNA) using the *WGCNA* package (v1.72.5), which identifies modules of co-expressed genes based on pairwise Pearson correlations in gene expression across samples^19^. Transcriptome-wide RNAseq profiles were filtered to retain genes with >5 counts in ≥1% of samples, normalized using variance stabilizing transformation^20^, and input to WGCNA to compute co-expression modules using a soft-thresholding power of 10. Each module was summarized by its module eigengene (first principal component) and assigned a color label for identification. Associations between variant carrier groups and co-expression modules were assessed using Pearson correlation between module eigengenes and binary carrier status vectors. Module-trait associations were retained if they met nominal significance (*p* < 0.05) and controls did not show a nominal association in the same direction. Putative associations were further assessed using multivariable linear regression, modeling module eigengenes as a function of variant carrier status while controlling for sex, ethnicity, batch effects, and variation in cellular composition; only associations with a Benjamini-Hochberg false discovery rate (FDR)-adjusted *p* < 0.05 were retained. Module hub genes were identified based on module membership (kME), defined as the correlation between each gene’s expression and the corresponding module eigengene. Genes were ranked by absolute kME, and the top 5% per module (minimum 25 genes) were retained as hub genes.

### Overrepresentation analysis

To characterize the biological functions represented by each co-expression module, we performed overrepresentation analysis (ORA) on module hub genes using the *gprofiler2* package (v0.2.3)^21^. ORA identifies gene subsets enriched within a target gene set relative to a background gene universe (all genes expressed in the dataset) using a cumulative hypergeometric test to evaluate whether the observed gene overlap exceeds chance expectation. Analyses were performed against the Gene Ontology (GO) Biological Process (BP), Molecular Function, and Cellular Component libraries, and terms passing FDR correction (*p* < 0.05) were considered significant^22^.

### Gene set variation analysis

To identify biological processes potentially driving co-expression module activity, we performed module-independent gene set variation analysis (GSVA) using the *GSVA* package (v1.46.0)^23^. GSVA is a sample-level enrichment method that transforms gene expression profiles into pathway activity scores by assessing coordinated expression of predefined gene sets across samples. Differential activity across variant carrier groups was evaluated using multivariable linear regression, modeling GSVA scores as a function of variant carrier status while controlling for sex, ethnicity, batch effects, and variation in cellular composition. Gene sets from the GO:BP library were queried, and terms with *p* < 0.01 were retained for downstream clustering.

To address the biological redundancy of the GO:BP library, terms were first clustered according to shared gene membership using pairwise Jaccard similarity and hierarchical clustering using average linkage and a dendrogram cut height of 0.95. Pairwise Pearson correlations were then calculated between cluster-level GSVA scores and clustered using Euclidean distance and complete linkage. The resulting groups, hereafter referred to as functional programs, represent higher-order biological units comprising overlapping GO:BP terms with correlated activity. Functional programs were manually annotated based on constituent terms and re-evaluated using GSVA, retaining programs passing FDR correction (*p* < 0.05). To minimize confounding in downstream analyses, only programs sharing <15% gene overlap with the corresponding co-expression module were retained.

### Structural equation modelling and Bayesian network analysis

To infer whether specific functional programs drive aberrant co-expression module activity in ALS variant carrier groups, we used structural equation modeling (SEM) to perform a series of complimentary mediation-style analyses with *semTools* (v0.5.7) and *lavaan* (v0.6.20).^24^ This framework estimated the effect of a genotype on module activity and the indirect effect transmitted through intermediate functional programs. A detailed description of the analytical framework is provided in **Supplementary File 1**.

To assess directional relationships between variants in ALS genes, functional program activity, and co-expression module activity, we first evaluated two competing mediation models. In Model 1, the effect of variants in ALS genes on co-expression module activity was hypothesized to be mediated by functional program activity. In Model 2, the ordering was reversed, testing whether functional program activity was mediated through co-expression module activity. Model fit was compared using Akaike Information Criterion (AIC)^25^. |Δ AIC| > 2 was used to support one model over the other, prioritizing functional programs that likely precede co-expression module perturbation in ALS variant carriers.

Next, we performed parallel mediation analyses using all functional programs with evidence of preceding co-expression module activity (Δ AIC > 2) as candidate mediators of the effect of variants in ALS genes on co-expression module activity. This framework estimates the mediating effects of each functional program simultaneously, while adjusting for all others. Functional programs showing significant indirect effects concordant with the observed effect of variant carrier status on co-expression module activity were retained.

To determine whether functional programs act in a sequential manner to influence co-expression module activity, we applied Bayesian network analysis. Bayesian networks were represented as directed acyclic graphs, in which nodes corresponded to variant carrier status, functional program activity, or co-expression module activity, while edges represented conditional dependencies between variables. Network structures were initially learned using score-based Bayesian network optimization and subsequently refined using automated heuristic structure search. To assess edge stability, bootstrap resampling was performed (*N* = 100 iterations). Edges detected in at least 50% of bootstrap iterations were retained as candidate pathways linking variants in ALS genes to co-expression module activity.

Finally, we evaluated all candidate pathways linking variants in ALS genes to co-expression module activity using serial mediation analysis to inform whether sequentially connected functional programs significantly mediated the effect of variants in ALS genes on co-expression module activity. Transcriptional pathways exhibiting significant serial effects after Bonferroni correction (*p* < 0.05) were subjected to genetic association analyses.

### Genetic association analysis

To investigate whether any genes identified in the transcriptional pathways showed variant- or gene-level associations with ALS, we queried summary statistics from large-scale genetic association studies. To assess potential impact of common variation, we used cross-ancestry genome-wide association study (GWAS) summary statistics from the analysis of 29,612 individuals with ALS and 122,656 controls^26^ and reported prioritized genes mapped to genome-wide significant loci (*p* < 5.00×10^-8^). To investigate potential impact from rare variation, we queried gene-burden summary statistics from the largest harmonized case-control exome sequencing based analysis in ALS to date, encompassing 17,919 individuals with ALS and 200,703 controls^27^. Specifically, variants were stratified according to (i) variant frequency, including ultrarare variants (URVs; variants observed in fewer than five individuals across the cohort) and singleton variants (variants observed only once across the dataset), and (ii) predicted functional impact, including high-impact variants alone (nonsense, splice acceptor/donor, and frameshift variants) or combined high- and moderate-impact variants (missense variants, in-frame deletions, and UTR truncations). Gene-level association statistics from the four variant categories were combined using aggregated Cauchy association tests (ACAT). We queried the presented results to identify genes reaching exome-wide significance after ACAT aggregation (17,324 genes; *p* < 2.89×10^-6^) as well as those surpassing Bonferroni correction within our prioritized gene set (2,599 genes; *p* < 1.92×10□□) in at least one variant category.

### Single-nuclei RNA sequencing analysis

To evaluate the cell type-specific expression of genes showing statistical associations with ALS, we analyzed a snRNAseq dataset profiling the motor and prefrontal cortices (MCx/FCx) of *C9orf72* ALS (C9ALS; *N* = 16), sporadic ALS (SALS; *N* = 17), and controls (*N* = 16)^28^. Processed data were retrieved from the *Synpase* portal (SynID: 51105515) and integrated into a unified *Seurat* (v5.3.1) object^29,30^. Cell type annotations from the original study were retained. Differential gene expression (DGE) between ALS and control samples was computed using *MAST* (v1.32.0), including sex as a latent variable. Genes with a |log2 fold-change (log2FC)| > 0.05 and FDR-adjusted *p* < 0.05 were considered differentially expressed^31^. To evaluate pathway-level activity, we computed GO:BP enrichment scores using the *UCell* package (v2.10.1) across all individual cells^32^. UCell scores were compared between ALS and control samples within each cell type and brain region using multivariable linear regression, modeling UCell scores as a function of disease status while controlling for sex. Gene sets with an FDR-adjusted *p* < 0.05 were considered differentially enriched.

### Statistical analysis

Statistical analyses were performed in R (v4.2.3) and data visualization was conducted using *ggplot2* (v4.0.1)^33,34^.

## Results

### Stratification of ALS subjects by variant carrier status

The Answer ALS dataset includes WGS data from PBMCs and bulk RNAseq data from iPSC-derived motor neurons. By integrating these datasets, individuals with ALS were stratified according to the presence of missense or pLoF variants in ALS genes (**Table 1**). Definitive ALS genes with >3 variant carriers were retained for downstream analyses, including *ANXA11* (*N_missense_ =* 4), *ATXN2* (*N_expansion_ =* 6), *C9orf72* (*N_expansion_ =* 30), *FUS* (*N_missense_ =* 3, *N_pLoF_* = 1), *GRN* (*N_missense_ =* 11), *NEK1* (*N_missense_ =* 7, *N_pLoF_* = 3), *OPTN* (*N_missense_ =* 2, *N_pLoF_* = 2), *SOD1* (*N_missense_* = 13), and *TBK1* (*N_missense_ =* 10, *N_pLoF_* = 1). ALS non-carriers (*N* = 265) and controls (*N* = 187) were retained as comparator groups.

**Table 1.**
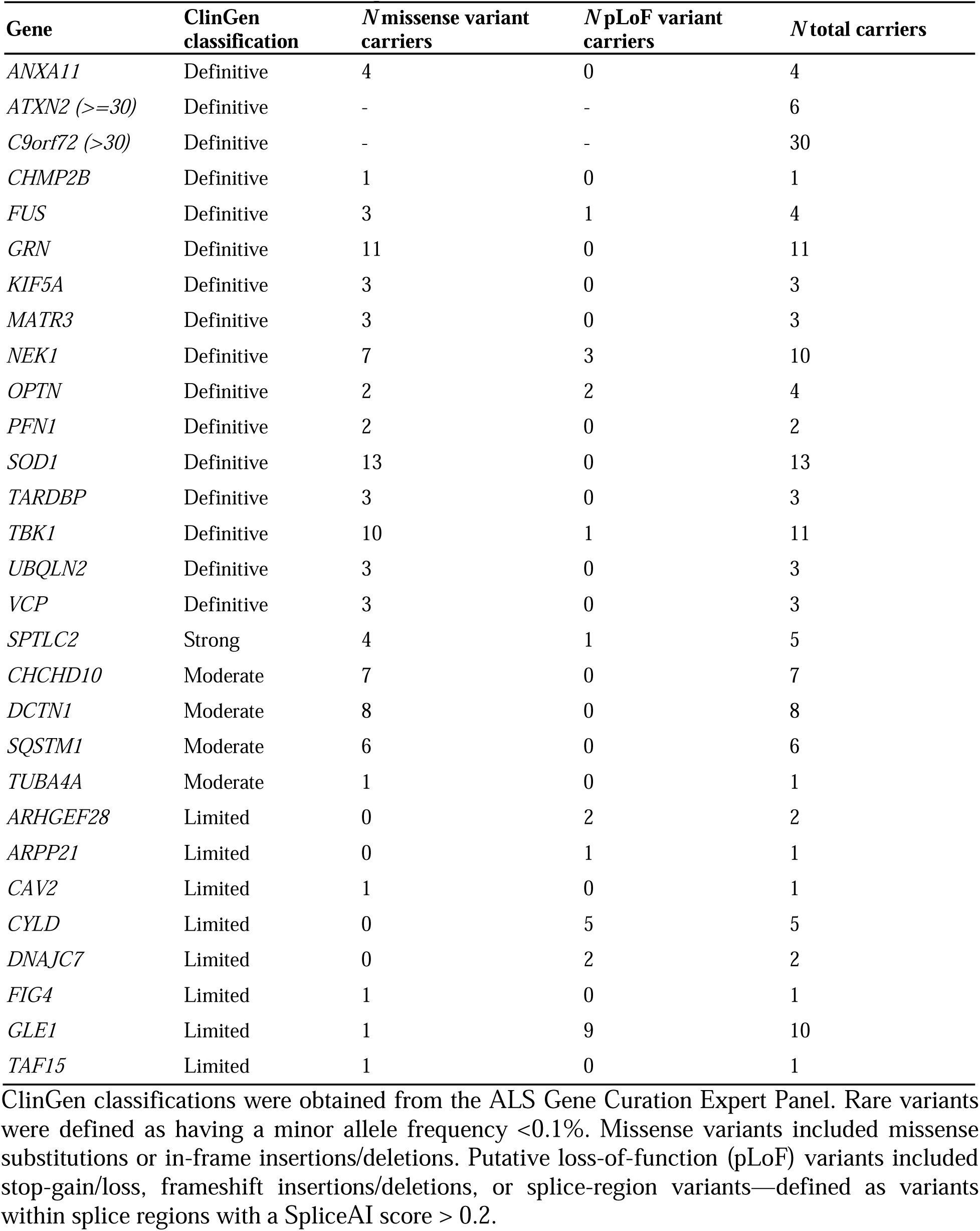
Individuals with ALS from the Answer ALS dataset (*N* = 438) that carried rare variants in known ALS associated genes.

We assessed whether individuals with ALS carrying non-synonymous variants in each definitive ALS gene exhibited altered expression of the mutated gene relative to ALS non-carriers and controls (**Supplementary Fig. 1A**). We observed significantly reduced expression of *C9orf72* in repeat expansion carriers relative to both ALS non-carriers (log2FC = -0.22, *p =* 3.28×10^-6^) and controls (log2FC = -0.34, *p* = 1.93×10^-9^). Consistent with this, snRNAseq analysis revealed reduced *C9orf72* expression across the majority of cell types in the MCx and FCx of individuals with C9ALS relative to SALS and controls (**Supplementary Fig. 1B**). Intriguingly, selectively vulnerable layer 5 (L5) neurons from the MCx exhibited the greatest reduction in *C9orf72* expression among all excitatory neuron subtypes. None of the other definitive ALS genes exhibited significantly altered expression in variant carriers.

### Network analysis identifies dysregulated co-expression modules across ALS variant carriers

To explore disease mechanisms independent of steady-state expression of the mutated gene, we performed WGCNA, which identified 26 modules of co-expressed genes representing coordinated transcriptional programs ranging in size from 35 to 2,860 genes. (**Supplementary Fig. 2**). Our two-step hurdle approach to assess differences in module activity across variant carrier groups revealed six significant module-trait associations (**Fig. 2A**).

**Fig. 2.**
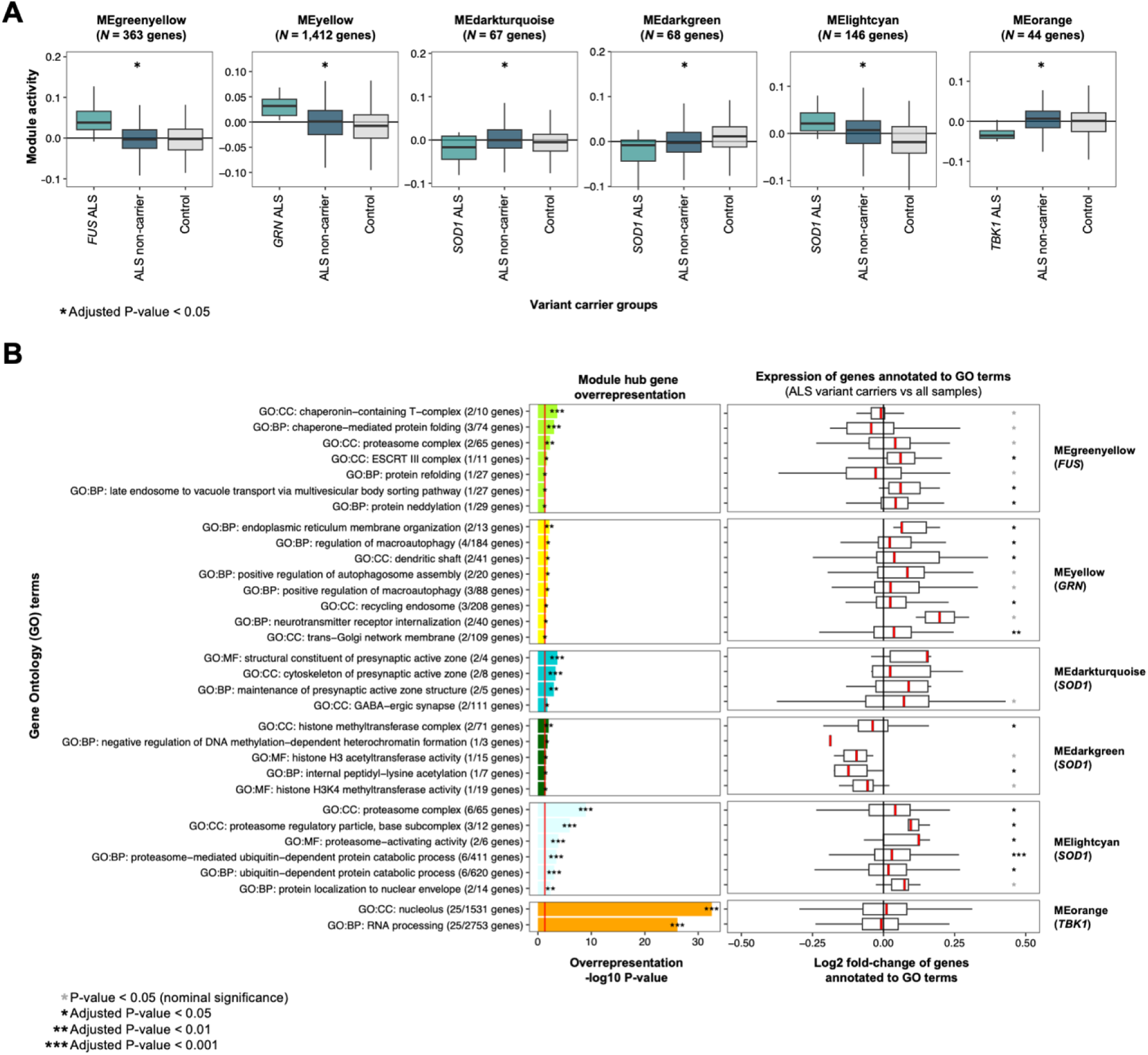
Co-expression modules with altered activity across genetic ALS subtypes. **A)** Weighted gene co-expression network analysis-derived module activity across ALS variant carrier groups. Differential module activity in variant carriers relative to all other samples wa assessed using multivariable linear regression. Associations with a Benjamini-Hochberg (BH) false discovery rate (FDR)-adjusted *p* < 0.05 are shown. **B)** Functional enrichment of module hub genes. Bar plots display the statistical significance (-log10 adjusted *P*-value) of Gene Ontology (GO) terms significantly enriched among module hub genes identified by overrepresentation analysis. Adjacent boxplots show the distribution of gene-level log2 fold-change values in expression between the corresponding ALS variant carrier group and all other samples for genes annotated to each functional term. A Wilcoxon signed-rank test was used to assess whether the distribution of log2 fold-change values differed significantly from 0. *P-*value were adjusted for multiple testing using the BH procedure to control the FDR.

Individuals with ALS carrying variants in *FUS* showed significantly increased activity of the MEgreenyellow co-expression module, which comprised 363 genes (β = 0.048, *p =* 8.20×10^-3^; **Fig. 2A**). The hub genes of this module were significantly enriched in proteostasis functions, including ATP-dependent and chaperone-mediated protein folding (*TCP1*, *CCT7*, *VBP1*, *PPIA*), proteasome function (*RAD23B*, *PSMG4*), prefoldin complexes (*TCP1*, *CCT7*, *VBP1*), ESCRT-III-mediated autophagy (*CHMP5*), and protein neddylation (*COPS4*), indicating a coordinated role in maintaining protein stability and mitigating cellular stress in *FUS*-associated ALS (**Fig. 2B; Supplementary Figs. 3A and 3B; Supplementary Table 1)**.

Individuals with ALS carrying variants in *GRN* showed significantly increased activity of MEyellow (1,412 genes; β = 0.024, *p =* 2.67×10^-2^; **Fig. 2A**). Hub genes were enriched for intracellular trafficking, including cytoskeletal transport (*STAU2, NSF, RAB6A, CDC42*), Golgi-to-plasma membrane trafficking (*RAB10, NSF, RAB6A, VTI1B, SCAMP1*), and endosomal pathways (*RTN4, RAB10, REEP5, RAB6A, SCAMP1, VTI1B*; **Fig. 2B; Supplementary Fig. 3C**). Hub genes also encompassed pathways related to autophagy (*PAFAH1B2, ATP6V1A, VTA1, RAB2A, SH3GLB1, MOAP1, VTI1B, CISD2*), ubiquitin ligase complexes (*KLHL9, KBTBD6, ZYG11B, KLHL8, KLHL7, UBE2V2*), and protein targeting to the endoplasmic reticulum and mitochondria (*RAB10, MOAP1, TOMM70, RTN4, VAPA, SH3GLB1, RALA*).

Individuals with ALS carrying *SOD1* variants showed significantly decreased activity of MEdarkgreen (68 genes; β = -0.023, *p =* 1.54×10^-2^) and MEdarkturquoise (67 genes; β = -0.022, *p* = 2.25×10^-2^), alongside significantly increased activity of MElightcyan (146 genes; β = 0.024, *p* = 1.46×10^-2^; **Fig. 2A**). MEdarkturquoise hub genes included *PCLO*, *BSN*, *EPB41L1*, *TANC2*, *NAV1*, *TRIM67*, *RAPH1*, *PACS1*, and *HUWE1*, implicating presynaptic active zone assembly, cytoskeletal organization, and synaptic vesicle transport (**Fig. 2B; Supplementary Fig. 3D**). MEdarkgreen hub genes were significantly enriched for pathways involved in histone lysine modifications (*EP300*, *KMT2A*, *BRD4*, *KMT2A*, *NCOA6*) and chromatin remodeling (*EP300*, *GATAD2B*, *KMT2A*, *SPEN*, *NFAT5*, *BRD4*, *BCORL1*, *UBN2*, *PHF21A*) (**Fig. 2B; Supplementary Fig. 3E**). MElightcyan defined a proteostasis network whose hub genes included *PSMB6*, *PSMB3*, *PSMB7*, *PSMD2*, *PSMC3*, and *PSMC4*, which are catalytic and regulatory subunits of the 26S proteasome, indicating activation of protein degradation programs **(Fig. 2B; Supplementary Fig. 3F).**

Individuals with ALS carrying variants in *TBK1* showed significantly decreased activity of MEorange (44 genes; β = -0.031, *p* = 3.30×10^-3^; **Fig. 2A**), whose hub genes consisted entirely of *SNORD115* family small nucleolar RNAs (snoRNA; **Supplementary Fig. 3G**). Collectively, the prominence of these snoRNAs implicated RNA processing as the primary biological function of the MEorange module **(Fig. 2B)**.

### Gene set variation analysis identifies perturbed biological processes across ALS variant carriers

Given that the co-expression modules were largely enriched for secondary or compensatory responses, we hypothesized that the observed transcriptional changes occur downstream of more proximal, gene-specific pathogenic effects. To investigate this, we performed GSVA to identify perturbed biological processes independent of the co-expression modules, including 425 significantly altered GO:BPs in *SOD1* ALS, 144 in *GRN* ALS, 133 in *TBK1* ALS, and 67 in *FUS* ALS (**Fig. 3A**). To reduce redundancy across significantly enriched GO:BPs, we clustered the processes based on gene overlap and correlated activity profiles, collapsing related processes into biologically coherent functional programs (**Fig. 3B; Supplementary Figs. 4-7**). This unsupervised clustering approach identified 26 functional programs in *SOD1* ALS, 27 in *GRN* ALS, 19 in *TBK1* ALS, and 15 in *FUS* ALS that remained significantly dysregulated following multiple hypothesis testing correction upon re-evaluation using GSVA (**Fig. 3C**).

**Fig. 3.**
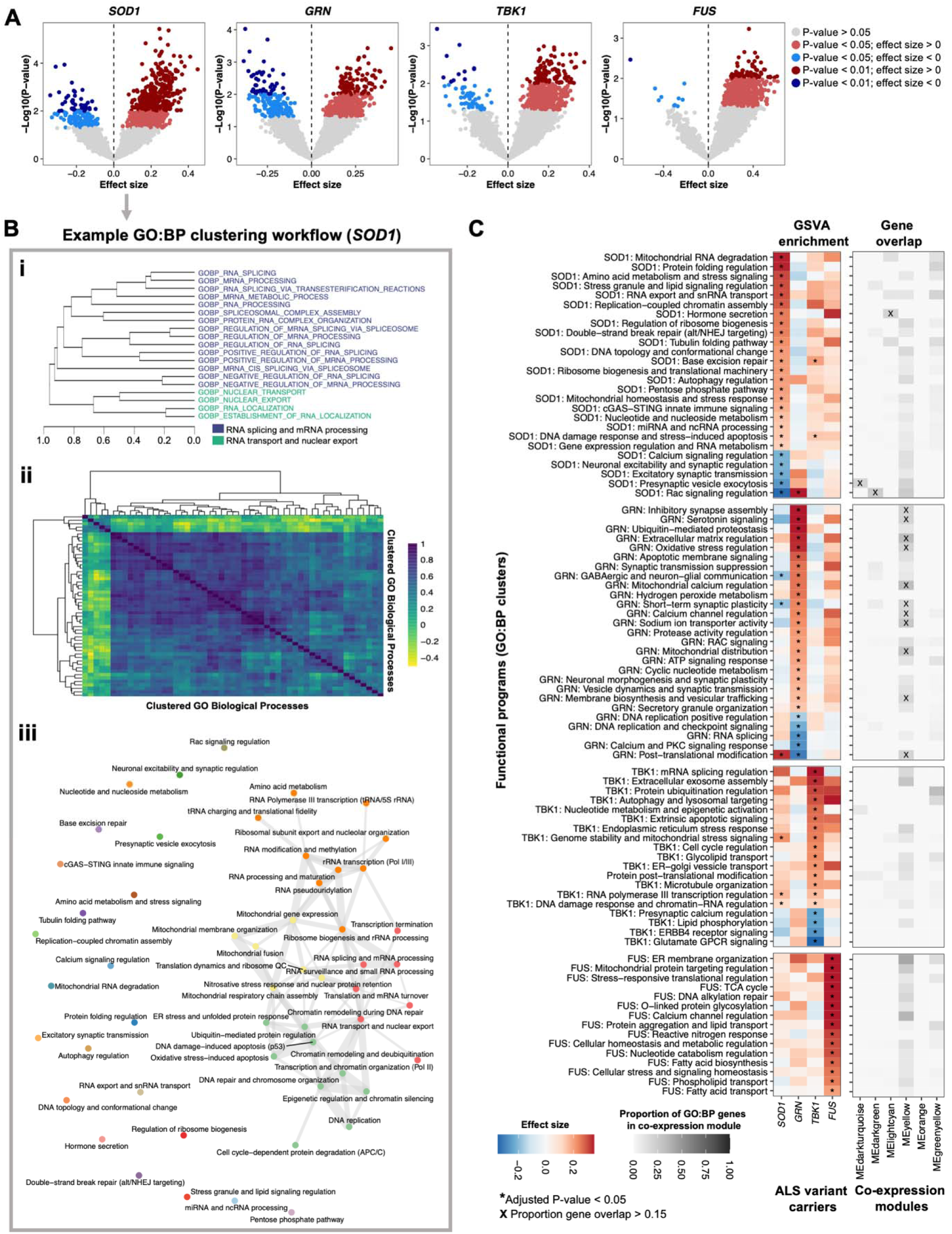
Functional programs with altered activity across genetic ALS subtypes. **A)** Volcano plots showing differential activity of Gene Ontology Biological Process (GO:BP) pathways quantified by gene set variation analysis (GSVA). Differential pathway activity in variant carriers relative to all other samples was assessed using multivariable linear regression. GO:BP terms with nominal significance (*P* < 0.01) were retained for downstream clustering analyses. **B)** Schematic overview of the GO:BP clustering workflow used to derive higher-order functional programs, shown using *SOD1* ALS as an example. **Bi)** Representative dendrogram illustrating hierarchical clustering of GO:BP terms based on shared gene membership. **Bii)** Heatmap showing pairwise Pearson correlations between cluster-level GSVA activity profiles. **Biii)** Network representation of functional programs derived from hierarchical clustering of the similarity matrix using Euclidean distance and complete linkage. Nodes represent GO:BP clusters, while edges represent correlation strength between clusters. **C)** Overview of the functional programs identified through the clustering workflow. The left heatmap shows differential functional program activity quantified by GSVA, with differential activity in variant carriers relative to all other samples assessed using multivariable linear regression. *P-*values were adjusted for multiple testing using the Benjamini-Hochberg procedure to control the false discovery rate. The right heatmap shows the proportion of genes within each functional program captured by the corresponding co-expression modules.

A subset of functional programs aligned with established primary pathogenic mechanisms of ALS genes (**Fig. 3C)**. *SOD1* ALS showed increased activity of *Mitochondrial homeostasis and stress response* (β = 0.19, *p* = 1.60×10^-3^), *Mitochondrial RNA degradation* (β = 0.34, *p* = 1.85×10^-3^), and *Protein folding regulation* (β = 0.34, *p* = 2.18×10^-4^), reflecting proteotoxic stress and mitochondrial dysfunction driven by mutant SOD1^35,36^. *GRN* ALS showed increased activity of *Membrane biosynthesis and vesicular trafficking* (β = 0.16, *p* = 2.56×10^-3^), *Secretory granule organization* (β = 0.15, *p* = 1.53×10^-3^), and *Vesicle dynamics and synaptic transmission* (β = 0.17, *p* = 6.13×10^-3^), consistent with progranulin-dependent defects in endolysosomal and secretory pathways^37^. *TBK1* ALS showed increased activity of *Autophagy and lysosomal targeting* (β = 0.30, *p* = 6.72×10^-3^) and *Protein ubiquitination regulation* (β = 0.31, *p* = 2.67×10^-2^), in line with TBK1’s central role in regulating autophagic flux and ubiquitin-mediated cargo recognition^38^. *FUS* variant carriers showed increased activity of *Stress-responsive translational regulation* (β = 0.35, *p* = 7.14×10^-3^) and *Protein aggregation and lipid transport* (β = 0.34, *p* = 2.88×10^-3^), reflecting disruption of RNA metabolism, stress granule dynamics, and proteostasis due to FUS mislocalization^39^.

### Structural equation modeling connects ALS genes to aberrant co-expression module activity

To infer whether specific functional programs drive aberrant co-expression activity across genetic ALS subtypes, we employed a multi-step framework using SEM and Bayesian networks. An example of this workflow for *SOD1* ALS is shown in **Fig. 4A**, with corresponding analyses for all ALS genes provided in **Supplementary Fig. 8**.

**Fig. 4.**
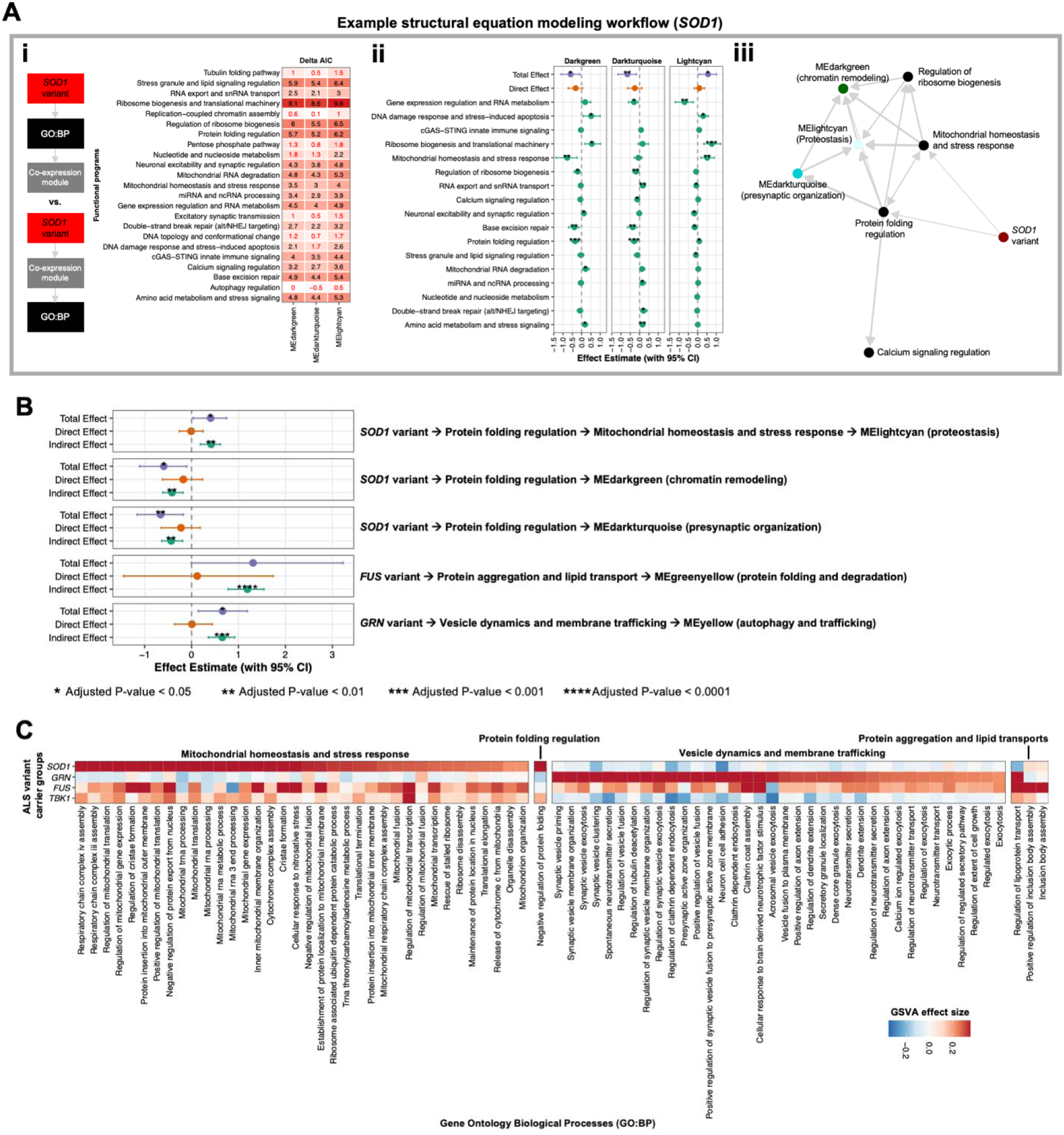
Transcriptional pathways linking variants in ALS genes to co-expression module activity. **A)** Overview of the analytical workflow used to nominate candidate transcriptional pathways linking variants in definitive ALS genes to altered co-expression module activity, shown using *SOD1* ALS as an example. **Ai)** Heatmap displaying ΔAIC values comparing two competing mediation models in which functional programs were modeled either as mediators of co-expression module activity or as downstream outcomes. Values are shown across all co-expression modules. Functional programs with ΔAIC > 2, indicating stronger support for the mediator model, were retained for downstream analyses. **Aii)** Total, direct, and indirect (mediated) effects estimated from parallel mediation models across all co-expression modules. Functional programs with significant indirect effects concordant with the observed effect of variant carrier status on co-expression module activity were retained for downstream analyses. Functional programs demonstrating significant inconsistent mediation for any module were excluded. **Aiii)** Network representation of the Bayesian network analysis used to infer directional relationships among variants in *SOD1*, functional programs, and co-expression modules. Nodes represent genetic status, functional programs, or co-expression modules, while edges represent inferred conditional dependency relationships between nodes. Candidate pathways identified from the Bayesian network were subsequently evaluated using serial mediation analysis. **B)** Total, direct, and indirect (mediated) effects estimated from serial mediation models. Only pathways meeting Bonferroni-adjusted significance (*p* < 0.05) are shown. **C)** Heatmap showing differential activity of the constituent Gene Ontology Biological Process terms comprising the mediating functional programs, quantified by GSVA.

The proposed framework identified candidate pathways linking variants in ALS genes to aberrant co-expression module activity through dysregulated functional programs (**Fig. 4B; Supplementary Table 2)**. The effects of *SOD1* variants on both the MEdarkgreen (chromatin remodeling) and MEdarkturquoise (presynaptic organization) co-expression modules were significantly mediated by the *Protein folding regulation* functional program (MEdarkgreen: *p* = 1.16×10^-4^; MEdarkturquoise: *p* = 1.11×10^-4^). Similarly, the effect of *SOD1* variants on MElightcyan (proteostasis) activity was significantly mediated by a serial path comprising the *Protein folding regulation* and *Mitochondrial homeostasis and stress response* functional programs (*p* = 1.06×10^-4^). The effect of *GRN* variants on MEyellow (autophagy and trafficking) activity was significantly mediated by the *Vesicle dynamics and membrane trafficking* functional program (*p* = 3.63×10^-6^). Finally, the effect of *FUS* variants on MEgreenyellow (protein folding and degradation) activity was significantly mediated by the *Protein aggregation and lipid transport* functional program (*p* = 8.14×10^-11^).

In **Fig. 4C**, we decomposed the mediating functional programs into their constituent GO:BP terms, revealing highly distinct transcriptional signatures across genetic ALS etiologies. *SOD1* variant carriers exhibited the strongest enrichment for processes related to mitochondrial homeostasis, with more moderate enrichment observed in *FUS*- and *TBK1*-associated ALS. Strong enrichment of processes related to proteostasis was unique to *SOD1* and *FUS* carriers, whereas GRN-associated ALS showed the strongest enrichment for pathways involved in vesicle dynamics and membrane trafficking.

### Transcriptional pathways implicate novel target ALS genes

To investigate whether genes comprising the empirically derived transcriptional pathways (*N* = 2,599 genes) exhibited genetic associations with ALS, we investigated summary statistics from large-scale genetic association studies. For common variant associations, we queried cross-ancestry GWAS summary statistics^26^. Of the 15 loci reaching genome-wide significance, five prioritized genes were captured within our transcriptional pathways (**Fig. 5A**). *SOD1* (rs80265967, European β = 1.08, European *p* = 3.5×10^-18^) mapped to the MEgreenyellow co-expression module, while *C9orf72* (rs2453555, cross-ancestry β = 0.168, cross-ancestry *p* = 1.5×10^-41^), *UNC13A* (rs12608932, European β = 0.125, European *p* = 8.8×10^-25^), *KIF5A* (rs113247976, European β = 0.332, European *p* = 1.4×10^-11^), and *PTPRN2* (rs10280711, cross-ancestry β = 0.086, cross-ancestry *p* = 1.8×10^-8^) were all implicated within the *Vesicle dynamics and membrane trafficking* functional program.

**Fig. 5.**
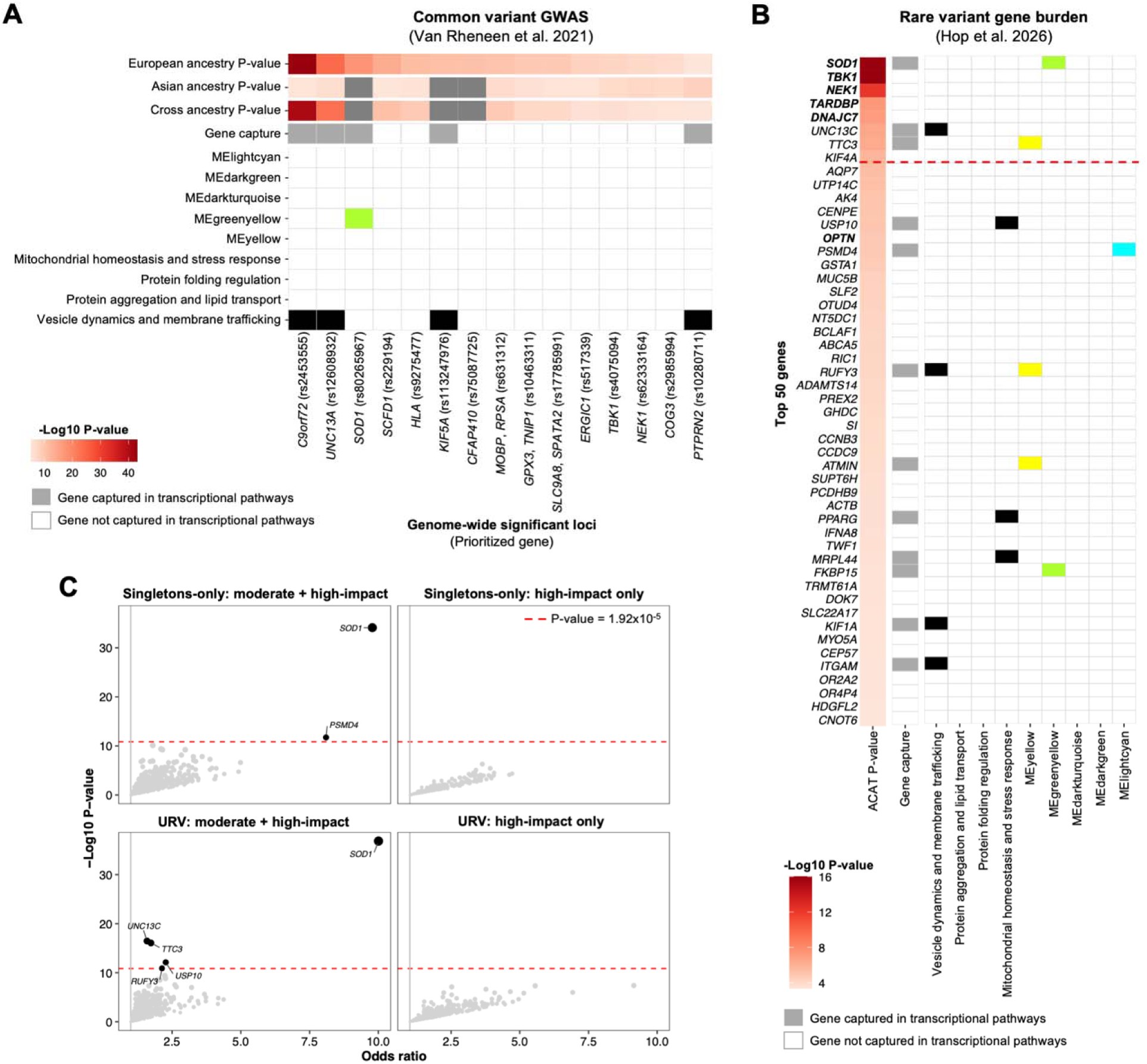
Prioritization of genetic targets embedded within transcriptional pathways associated with definitive ALS genes. **A)** Heatmap showing prioritized genes at genome-wide significant loci identified in the most recent cross-ancestry ALS genome-wide association study (GWAS), encompassing 29,612 individuals with ALS and 122,656 controls. Genes mapped to genome-wide significant loci (*p* < 5.00×10^-8^) are shown, with those represented within the identified transcriptional pathways highlighted. **B)** Heatmap showing the top 50 most significant genes from the largest rare variant gene-burden analysis in ALS to date, encompassing 17,919 individuals with ALS and 200,703 controls. *P* values correspond to aggregated Cauchy association tests (ACAT), which combine gene-level association statistics across the four variant categories shown in panel C. The red dashed line indicates the Bonferroni-corrected significance threshold for 17,324 protein-coding genes (*p* < 2.89×10^-6^). Genes captured by transcriptional pathways are highlighted. **C)** Scatter plots showing gene-level association statistics from rare variant burden analyses across four variant categories. The red dashed line indicates the Bonferroni-corrected significance threshold accounting for the size of the prioritized gene set (*N* = 2,599; *p* < 1.92×10□□). Genes surpassing this threshold are labeled.

For rare variant associations, we queried rare variant gene-burden summary statistics across four variant categories, as well as ACAT-based aggregation across all categories^27^. Among the 50 most significantly enriched genes in individuals with ALS following ACAT aggregation, 12 were captured within our transcriptional pathways (**Fig. 5B**), representing a significant enrichment relative to chance expectation (*p* = 4.00×10□²; odds ratio [OR] = 1.94). Across 17,324 protein-coding genes analyzed in the original rare variant analysis, eight reached exome-wide significance, including three emerging target genes: *TTC3* (*p* = 4.16×10□□), *UNC13C* (*p* = 2.80×10□□), and *KIF4A* (*p* = 1.62×10□□) (**Fig. 5B**). Remarkably, our transcriptional pathways captured two of three emerging targets: *UNC13C* was implicated in the *Vesicle dynamics and membrane trafficking* functional program, while *TTC3* was a member of the MEyellow co-expression module.

To nominate novel target genes associated with ALS, we applied a Bonferroni correction adjusted for the size of our prioritized gene set (2,599 genes; *p* < 1.92×10□□) to the rare variant gene-burden analysis. In doing so, we identified three additional associations: *USP10* (OR = 2.27, *p* = 5.40×10^-6^) and *RUFY3* (OR = 2.13, *p* = 1.83×10^-5^) which showed enrichment of URVs with moderate- or high-impact consequences in individuals with ALS, and *PSMD4* (OR = 8.10, *p* = 8.02×10^-6^), which showed enrichment of singleton variants with moderate- or high-impact consequences in individuals with ALS (**Fig. 5C**). *USP10* was implicated in the *Mitochondrial homeostasis and stress response functional program*, while *RUFY3* and *PSMD4* were members of the Meyellow and MElightcyan co-expression modules, respectively.

### ALS target genes are enriched in the intrinsic architecture of neuronal populations

Collectively, the transcriptional pathways captured 10 genes with statistically significant associations with ALS based on previously published genetic studies. These included established ALS genes (*C9orf72*, *UNC13A*, *SOD1*, and *KIF5A*), three emerging targets that have been previously implicated in ALS (*PTPRN2*, *UNC13C*, and *TTC3*), and three novel targets (*USP10*, *PSMD4*, and *RUFY3*). Leveraging snRNAseq, we examined the cell type-specific expression of these genes in the MCx and FCx of neurologically healthy controls, revealing predominant expression in neuronal cell types across both cortical regions, relative to glial and vascular populations (**Fig. 6A**). This pattern remained consistent following further stratification into transcriptionally defined subtypes (**Supplementary Fig. 9**). To quantify this enrichment, we assessed the aggregate expression of ALS genes across major cell type classes (neuronal, glial, and vascular) and within neuronal subclasses (excitatory neurons, inhibitory neurons, and non-neuronal populations). Neuronal populations were significantly enriched for expression of established, emerging, and novel ALS genes compared with glial and vascular populations (**Fig. 6B**). Similarly, excitatory neurons showed significantly higher enrichment across all gene categories relative to inhibitory neurons and non-neuronal populations.

**Fig 6.**
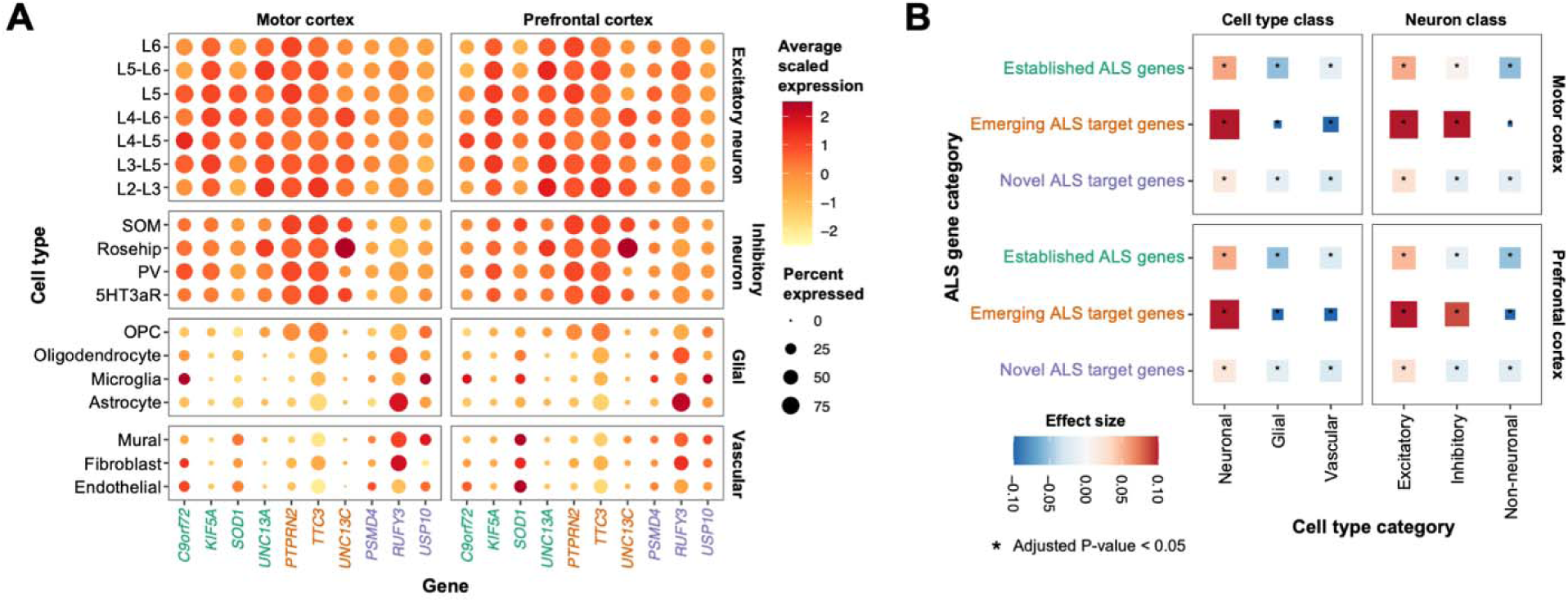
Cell type enrichment of ALS target genes across cellular populations in the motor and prefrontal cortices. The single-nucleus RNA sequencing data are derived from the motor and prefrontal cortices (MCx/FCx) of neurologically healthy controls. **A)** Dotplot showing the average scaled expression and percent expression of established ALS genes (green), emerging ALS target genes (orange), and novel ALS target genes (purple) across major cell types comprising the MCx and FCx. **B)** Dot plot showing differential enrichment of ALS gene categories across cell type and neuronal classes, stratified by brain region. Cumulative ALS gene expression was quantified using *UCell*, and differential enrichment was assessed using multivariable linear regression across cell type and neuron class strata. Point size reflects the effect size of the regression model. *P-*values were adjusted for multiple testing using the Benjamini-Hochberg procedure to control the false discovery rate.

### *PSMD4* dysregulation implicates region-specific stress programs in vulnerable L5 motor neurons

To investigate neuron-intrinsic pathological mechanisms of target genes showing significant genetic associations with ALS, we performed DGE analyses between ALS and controls using bulk RNAseq of iPSC-derived motor neurons and snRNAseq of post-mortem MCx and FCx tissue. Among emerging target genes, ALS non-carriers exhibited increased expression of both *TTC3* (log2FC = 0.01, *p =* 6.62×10^-3^) and *PTPRN2* (log2FC = 0.12, *p =* 1.66×10^-5^) relative to controls in iPSC-derived motor neurons (**Supplementary Fig. 10A**). In post-mortem tissue, *PTPRN2* dysregulation was broadly replicated, showing significantly increased expression across neuronal populations in both SALS cortices, whereas *TTC3* expression was not significantly altered (**Supplementary Fig. 10B)**. *UNC13C* did not show consistent differential expression in either the bulk or single-cell datasets.

Among novel target genes, ALS non-carriers showed moderately reduced expression of *USP10* relative to controls in iPSC-derived motor neurons (log2FC = 0.02, *p =* 6.62×10^-3^); however, *USP10* expression was significantly increased across the majority of neuronal populations in the post-mortem SALS cortices (**Supplementary Figs. 10A and 10B**). In contrast, *RUFY3* exhibited widespread upregulation in iPSC-derived motor neurons across multiple genetic ALS subtypes, including *C9orf72* (log2FC = 0.05, *p =* 1.15×10^-2^), *GRN* (log2FC = 0.13, *p =* 2.45×10^-3^), *NEK1* (log2FC = 0.11, *p =* 6.62×10^-3^), and *SOD1* ALS (log2FC = 0.10, *p =* 2.52×10^-2^), but this pattern was not observed in neuronal populations from the post-mortem SALS cortices.

Most notably, the iPSC-derived motor neurons revealed significant dysregulation of the novel target gene *PSMD4*. Expression was increased in both *SOD1* ALS (log2FC = 0.21 *p* = 1.52×10^-4^) and ALS non-carriers relative to controls (log2FC = 0.07, *p* = 1.87×10^-5^) (**Fig. 7A**). At single-cell resolution, increased *PSMD4* expression in SALS excitatory neurons was largely restricted to selectively vulnerable L5 motor neurons of the post-mortem MCx (log2FC = 0.19, *p* =1.38×10^-8^) (**Fig. 7B; Supplementary Fig. 10B**). This pattern persisted following refinement of excitatory neurons into transcriptionally defined subtypes, with the strongest differential expression observed in the L5 *VAT1L THSD4* subtype (log2FC = 0.37, *p* = 6.57×10^-3^). In contrast, this L5-specific upregulation was not observed in the SALS FCx, where *PSMD4* expression instead trended toward downregulation, although the effect did not reach statistical significance (log2FC = -0.12, *p* =4.73×10^-1^) (**Fig. 7B)**.

**Fig. 7.**
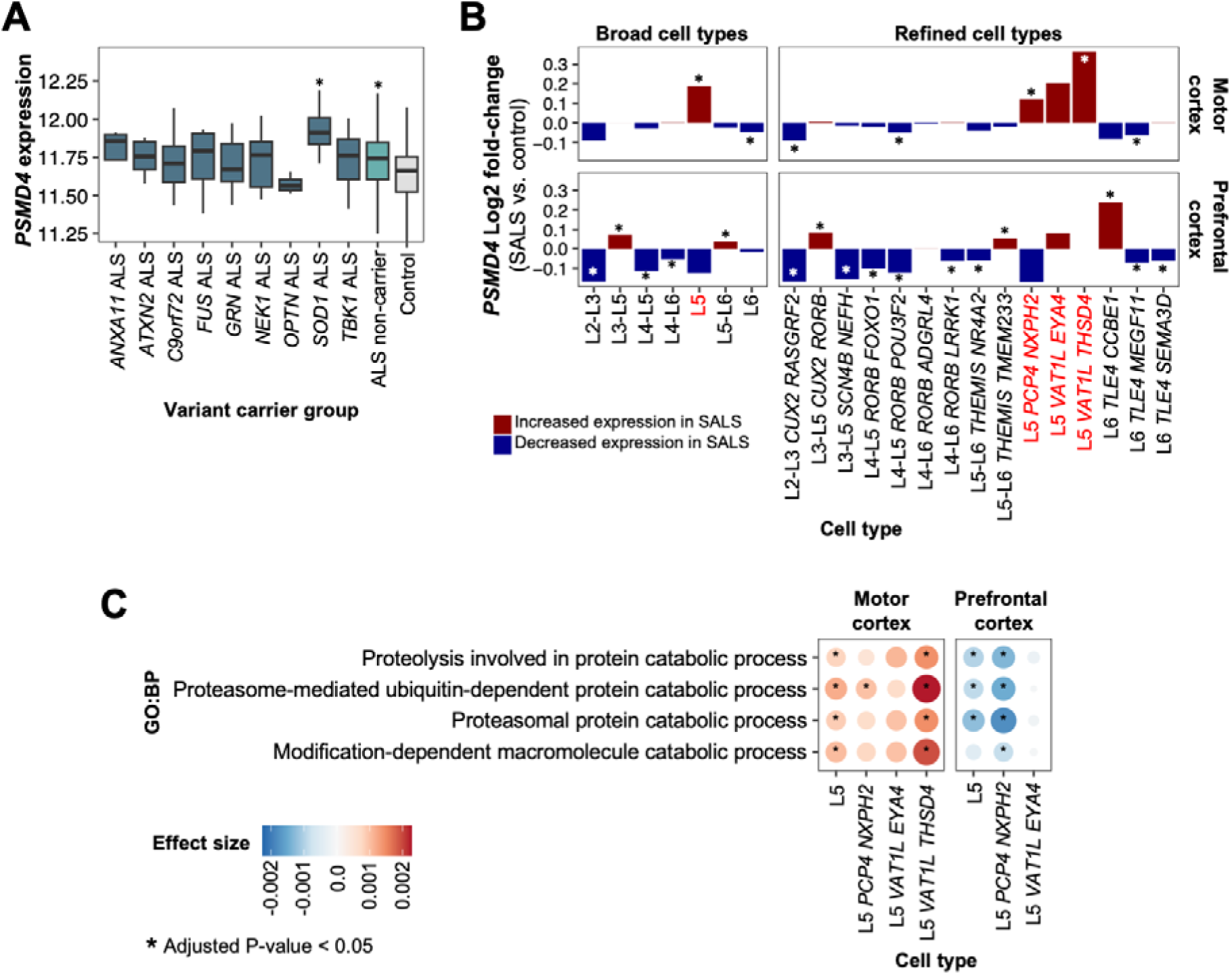
Region-specific differential expression of *PSMD4* in Layer 5 motor neurons. **A)** *PSMD4* expression across variant carrier groups in bulk RNAseq data from induced pluripotent stem cell-derived motor neurons. Differential expression between ALS variant carriers and controls was assessed using a Wilcoxon rank-sum test. **B-C**) The single-nucleus RNA sequencing data are derived from the motor and prefrontal cortices (MCx/FCx) of individual with sporadic ALS (SALS) neurologically healthy controls. **B)** Differential expression of *PSMD4* in excitatory neurons between SALS and controls, stratified by brain region. Differential expression was computed using MAST. **C)** Dot plot showing differential activity of Gene Ontology Biological Process (GO:BP) pathways containing *PSMD4* across Layer 5 motor neuron subtypes, stratified by brain region and quantified using *UCell*. Differential pathway activity between SALS and controls was assessed using multivariable linear regression. *P-*value presented in panels A-C were adjusted for multiple testing using the Benjamini-Hochberg procedure to control the false discovery rate.

To determine whether *PSMD4* dysregulation reflected broader perturbations in proteostasis-related stress programs, we evaluated pathway-level transcriptional activity for all GO:BPs containing *PSMD4*, including *Proteolysis involved in protein catabolic process*, *Proteasome-mediated ubiquitin-dependent protein catabolic process*, *Proteasomal protein catabolic process*, and *Modification-dependent macromolecule catabolic process* (**Fig. 7C**). Across these proteostasis-related pathways, we observed increased activity in SALS L5 motor neurons of the MCx relative to controls, whereas the same neuronal populations in the FCx exhibited decreased pathway activity in SALS relative to controls, reflecting the gene-level expression patterns of *PSMD4*.

## Discussion

Here, we leveraged an integrative framework to model molecular perturbations downstream of variants in definitive ALS genes and investigated whether disease-associated transcriptional pathways harbor novel genetic risk factors. The analysis characterized the molecular consequences of genetic ALS subtypes and identified novel target disease genes, offering mechanistic insight into their potential roles in disease pathogenesis.

Consistent with prior studies^40,41^, individuals with ALS carrying *C9orf72* repeat expansions exhibited reduced *C9orf72* expression relative to both ALS non-carriers and controls. Extending beyond previous findings, our analyses demonstrated that this reduction was most pronounced in L5 neurons of the MCx, a neuronal population selectively vulnerable to degeneration in ALS^42,43^. Given that reduced *C9orf72* expression impairs autophagy and the clearance of toxic dipeptide repeat proteins^44^, the reduction in *C9orf72* expression observed in L5 neurons may exacerbate proteostatic dysfunction in a cell type already burdened by high metabolic demands and extensive long-range axonal projections^45^, thereby contributing to its selective vulnerability in ALS.

Individuals with ALS carrying *FUS* variants showed increased activity of a co-expression module enriched for protein folding and degradation pathways. Specifically, pathways involved in prefoldin-mediated protein folding, protein neddylation, and ESCRT-III-mediated autophagy were enriched, delineating the specific proteostasis pathways engaged in this genetic ALS subtype. We further found that the effect of *FUS* variants on co-expression module activity was significantly mediated by the *Protein aggregation and lipid transport* functional program, consistent with the tendency of mutant FUS to form aggregates^46,47^ and disrupt protein quality-control pathways^48^. While aberrant lipid metabolism in response to mutant FUS has been described in skeletal muscle of *FUS*-associated ALS mouse models^49^, our findings extend this process to the ALS brain, where it emerges as a potential early event coinciding with proteostatic stress. Notably, dysregulated lipid homeostasis has been shown to disrupt ESCRT function, a primary component of the co-expression module, and impair proteostatsis^50^. Collectively, our results support a model in which aberrant protein aggregation and disrupted lipid handling converge to drive proteostasis stress in *FUS* ALS.

Individuals with ALS carrying variants in *GRN*, which encodes progranulin, exhibited increased activity of a co-expression module enriched for intracellular trafficking, including Golgi-to-plasma membrane and endosomal transport, as well as autophagy. These findings align with evidence that progranulin deficiency disrupts endolysosomal trafficking, biasing trafficking decisions away from lysosomal pathways toward extracellular vesicle release^37^ and promoting the intercellular spread of pathogenic proteins^51–53^. The effect of *GRN* variants on module activity was significantly mediated by the *Vesicle dynamics and membrane trafficking* functional program, reflecting a coordinated reconfiguration of endosomal sorting and vesicular transport in response to impaired lysosomal degradation capacity. Furthermore, processes related to synaptic maintenance co-occurred with this functional program, suggesting that trafficking defects may impair synaptic connectivity and plasticity through altered endosomal recycling of synaptic membrane components in progranulin deficiency. Collectively, these results suggest that trafficking rerouting is a central mechanistic feature of *GRN*-associated ALS, linking endolysosomal dysfunction to both proteostatic stress and synaptic dysfunction.

Individuals with ALS carrying *SOD1* variants exhibited reduced activity of two co-expression modules enriched for synaptic processes and chromatin remodeling, respectively. Both effects were significantly mediated by the *Protein folding regulation* functional program, implicating proteostatic imbalance as a central driver of the transcriptional changes. While mutant SOD1 has been shown to disrupt mitochondrial dynamics, resulting in the mislocalization of mitochondria at motor neuron synapses and consequent impairment of synaptic function^54^, our findings suggest that SOD1 aggregation may also compromise synaptic integrity independently through defective protein quality control. Impaired protein folding could disrupt the folding, trafficking, and turnover of synaptic and cytoskeletal proteins required to maintain synaptic architecture and neurotransmission, providing a complementary mechanism by which mutant SOD1 promotes synaptic dysfunction. Likewise, the mediation of reduced chromatin remodeling activity by the *Protein folding regulation* program suggests that the consequences of SOD1 misfolding extend beyond cytoplasmic proteostasis to influence nuclear regulatory processes. This is consistent with evidence that mutant SOD1 induces oxidative stress, DNA damage, and epigenetic alterations that collectively remodel chromatin accessibility and transcriptional programs in ALS^55^.

In parallel, ALS *SOD1* variant carriers exhibited increased activity of a co-expression module that defined a proteostasis network whose activity was serially mediated by the *Protein folding regulation* and *Mitochondrial homeostasis and stress response* functional programs. This finding aligns with evidence that misfolded SOD1 accumulations impair mitochondrial function^56^ and supports a feed-forward model in which protein aggregation and mitochondrial dysfunction reciprocally reinforce one another, amplifying proteostatic stress through impaired bioenergetics and oxidative stress. Collectively, these findings suggest that *SOD1*-associated ALS is characterized by coordinated disruption of synaptic and nuclear regulatory programs alongside activation of compensatory proteostasis pathways.

The transcriptional pathways identified in this study encompassed 10 genes showing significant genetic associations with ALS, including three emerging targets in ALS (*PTPRN2*, *UNC13C*, and *TTC3*)^26,27^, and three novel targets (*USP10*, *PSMD4*, and *RUFY3*) that, to our knowledge, have not previously been reported in the context of ALS genetic association studies. Indeed, these pathways were significantly enriched for the top 50 most significant rare variant gene-burden signals, supporting the central premise of this work that disease-associated transcriptional perturbations can be leveraged to prioritize candidate genes for genetic discovery. This is particularly important in ALS, where a substantial proportion of genetic risk is driven by rare variants^57^, limiting detection by genome- and exome-wide approaches. We anticipate that this framework will prove broadly applicable for the discovery of elusive genetic risk factors across complex heritable diseases.

The genes identified in this study with significant genetic associations to ALS were preferentially expressed in neurons relative to glial and vascular populations. These findings are consistent with reports demonstrating neuronal enrichment of ALS risk loci but extend previous work by resolving enrichment to excitatory neuronal populations^26^. This pattern was observed for both established ALS genes and the novel target genes, providing independent support for their relevance to ALS pathobiology. Importantly, the enrichment was observed in neurologically healthy individuals, indicating that elevated expression of ALS risk genes reflects intrinsic neuronal transcriptional programs rather than disease-induced activation. These observations suggest that ALS genetic risk is embedded within the molecular architecture of excitatory neurons, supporting a model in which neurodegeneration is driven primarily by neuron-intrinsic mechanisms.

The functions of the novel targets identified in this study further support a role for neuron-intrinsic mechanisms in ALS pathogenesis. *USP10* encodes a deubiquitinating enzyme involved in ubiquitin signaling and cellular stress responses, with established roles in regulating p53 stability, stress granule dynamics, and autophagy-related protein quality control, processes central to neuronal proteostasis^58^. *USP10* expression was increased across most neuronal populations in post-mortem SALS cortices relative to controls, which may reflect an adaptive response to chronic proteostatic stress and misfolded protein burden. In this context, we can propose that genetic variation in *USP10* may further modulate deubiquitinating activity and stress-response signaling. Subtle perturbations in these pathways could reduce neuronal capacity to maintain proteostasis under chronic stress conditions, thereby increasing vulnerability to dysfunction in ALS.

*RUFY3* is a regulator of axonal growth and endolysosomal trafficking that supports retrograde transport and the maintenance of neuronal processes^59^, pathways that are increasingly implicated in motor neuron degeneration^60^. We observed increased *RUFY3* expression across multiple genetic ALS etiologies in iPSC disease models relative to controls; however, this pattern was not observed in post-mortem SALS cortices. Further work is needed to establish the transcriptional patterns of *RUFY3* in ALS. Nonetheless, it is tempting to speculate that deleterious *RUFY3* variants could contribute to ALS risk by conferring progressive deficits in cargo clearance and intracellular homeostasis, which may be particularly relevant in long-projection motor neurons that are known to be sensitive to even modest trafficking impairments^61^.

Finally, *PSMD4* functions as a ubiquitin receptor within the 26S proteasome and is required for ubiquitin-mediated protein degradation^62^. We observed region-specific alterations in *PSMD4* expression within vulnerable L5 neurons, with increased expression in the SALS MCx and decreased expression in the FCx compared to controls. The strongest increase occurred in the L5 *VAT1L THSD4* subtype, an MCx-specific population enriched for ALS GWAS-associated genes and marked transcriptional divergence in ALS^28^. These changes paralleled regional differences in proteasome activity, which was elevated in L5 neurons of the SALS MCx and reduced in the FCx relative to baseline. Increased activity in MCx may reflect a compensatory response to proteotoxic stress, whereas reduced activity in FCx could indicate exhaustion of proteostatic mechanisms during later stages of degeneration, consistent with the progressive loss of FCx L5 projection neurons in ALS^63^. Together, these findings implicate regionally divergent proteasome activation as a neuron-intrinsic feature of ALS vulnerability, potentially driven in part by dysregulation of *PSMD4*.

We also recognize the limitations of our work. For example, the stratification of ALS samples by variant carrier status resulted in small sample sizes for several definitive ALS genes, limiting statistical power and complicating the interpretation of negative findings. Further, the mediation analyses reflect statistical dependencies inferred from cross-sectional transcriptomic data and therefore cannot distinguish causal drivers from correlated or downstream responses within complex regulatory networks. Nevertheless, the strong concordance between the inferred pathways and established ALS mechanisms supports the biological relevance of this approach. Finally, these findings are based solely on transcriptomic data and lack experimental validation. Future functional studies will be required to establish the mechanistic relevance of the identified pathways and clarify the potential contribution of the novel target genes to ALS pathogenesis.

In conclusion, our integrative analysis characterized the downstream molecular consequences of variants in definitive ALS genes and illustrated how transcriptomic signatures can be leveraged to identify biologically relevant gene sets to enhance the discovery of genetic risk factors. Our findings provide additional support for the involvement of the emerging ALS target genes *PTPRN2*, *UNC13C*, and *TTC3*, while nominating *USP10*, *PSMD4*, and *RUFY3* as novel targets. Together, these results offer new insight into the molecular pathways linking established ALS genetics to downstream disease mechanisms and underscore the broader potential of integrative multi-omic approaches to advance our understanding of complex neurodegenerative diseases.

## Abbreviations

ALS: amyotrophic lateral sclerosis
AIC: Akaike Information Criterion
DAG: directed acyclic graphs
DGE: differential gene expression
FDR: false discovery rate
GCEP: Gene Curation Expert Panel
GO:BP: Gene Ontology Biological Process
GSVA: gene set variation analysis
GWAS: genome-wide association study
iPSC: induced pluripotent-stem cell
kME: module membership
L5: layer 5
MAF: minor allele frequency
OR: odds ratio
ORA: overrepresentation analysis
PBMC: peripheral blood mononuclear cell
pLoF: putative loss-of-function
RNAseq: RNA sequencing
scRNAseq: SEM: structural equation modelling
snoRNA: small nucleolar RNA
snRNAseq: single-nuclei RNA sequencing
URV: ultra-rare variant
VEP: Variant Effect Predictor
WGCNA: weighted gene co-expression network analysis
WGS: whole-genome sequencing.

## Data availability

Bulk RNAseq data from iPSC-derived motor neurons and WGS data from PBMCs were obtained from the Answer ALS Data Portal and are available upon registration. SnRNAseq data generated by Pineda et al., are available through the Synapse data repository under accession number 51105515 (https://www.synapse.org/). The code used to for the analyses presented in this study is available on GitHub (https://github.com/mfiorini9/AALS-analysis).

## Acknowledgments

Data used in the preparation of this article were obtained from the ANSWER ALS Data Portal (**AALS-01184**). For up-to-date information on the study, visit https://dataportal.answerals.org

## Funding

MRF was supported by a CIHR Canada Graduate Scholarships-Master’s Award, a Fonds de Recherche Santé Québec Master’s Award, and a CIHR Canada Graduate Scholarships-Doctoral Award (536860). SMKF received funding from Brain Canada and the Montreal Neurological Institute-Hospital.

## Competing interests

The authors report no competing interests.

## Supplementary material

**Supplementary File 1:** Supplementary Methods.

**Supplementary File 2.** Supplementary Figures 1-10.

**Supplementary Table 1.** Functional overrepresentation analysis of co-expression module hub genes.

**Supplementary Table 2.** Statistical results of the serial mediation analyses.

## Notes

### Competing Interest Statement

The authors have declared no competing interest.

### Author Declarations

The study used only openly available data that were originally located at the Answer ALS Data Portal and Synapse.

