## Supplementary File 1 for "Transcriptional pathways of definitive ALS genes implicate novel disease-associated target genes"

**Supplementary Methods**

**Structural equation modelling and Bayesian network analysis**

To infer whether specific functional programs drive aberrant co-expression module activity in ALS variant carrier groups, we used structural equation modeling (SEM) to perform a series of complimentary mediation-style analyses with *semTools* (v0.5.7) and *lavaan* (v0.6.20). This framework estimated the effect of a genotype on module activity and the indirect effect transmitted through intermediated functional programs. All SEMs were fit using maximum likelihood estimation with 5,000 bootstrap iterations and included sex, ethnicity, batch effects, and variation in cellular composition as covariates. We restricted this analysis to ALS samples to specifically assess the effects of variants.

To infer directional relationships between variants in ALS genes, functional program activity, and co-expression module activity, we evaluated two competing mediation models. In Model 1, the effect of variants in ALS genes on co-expression module activity was hypothesized to be mediated by functional program activity:

$$Functional program activity=a\cdot{Variant}_{carrier}+C\beta+ℇ_{1}$$

$$Coexpression module activity=b\cdot Functional program activity+C\gamma+ℇ_{2}$$

Where $Functional program activity$ represents the GSVA score of the functional program, $Coexpression module activity$ represents the co-expression module’s eigengene, ${Variant}_{carrier}$ denotes binary variant carrier status for a given ALS gene, and $C$ represents covariates. The indirect effect was estimated as:

$$Indirect effect=a\times b$$

In model 2, the ordering was reversed, testing whether functional program activity was mediated through co-expression module activity:

$$Coexpression module activity=a\cdot{Variant}_{carrier}+C\beta+ℇ_{1}$$

$$Functional program activity=b\cdot Coexpression module activity+C\gamma+ℇ_{2}$$

with the indirect effect similarly defined as $a\times b$. Model fit was compared using Akaike Information Criterion (AIC), with lower values indicating improved model support. |Δ AIC| > 2 was used to support one model over the other, thereby distinguishing functional programs that likely precede versus follow co-expression module perturbation in ALS variant carriers.

Next, we performed parallel mediation analyses using all functional programs with evidence of preceding co-expression module activity (Δ AIC > 2) as candidate mediators of the effect of variants in ALS genes on co-expression module activity. This framework estimates the mediating effects of each functional program simultaneously, while adjusting for all others, thereby prioritizing functional programs that contribute to co-expression module activity rather than reflecting shared or correlated activity. For each mediator $M_{j}$, the following regression was fit:

$$M_{j}= a_{j}\cdot{Variant}_{carrier}+C\beta_{j}+ℇ_{j}$$

where $M_{j}$ represents the GSVA score of the functional program $j$. Co-expression module activity was then modeled as:

$$Coexpression module activity=c^{'}\cdot{Variant}_{carrier}+\sum_{j=1}^{k} b_{j}M_{j}+C\gamma+ℇ$$

where $c^{'}$ represents the direct effect of variant carrier status on module activity after accounting for all mediators, and $b_{j}$ represents the effect of mediator $M_{j}$ on module activity. The indirect effect mediated through each functional program was defined as:

$${Indirect}_{j}=a_{j}\times b_{j}$$

and the total indirect effect across all mediators was defined as:

$$Total indirect effect= \sum_{j=1}^{k} (a_{j}\times b_{j})$$

The total effect of variant carrier status on module activity was defined as:

$$Total effect=c^{'}+\sum_{j=1}^{k} (a_{j}\times b_{j})$$

Functional programs showing significant indirect effects concordant with the observed effect of variant carrier status on co-expression module activity were retained for downstream analyses. For *SOD1* ALS, which exhibited dysregulation across multiple co-expression modules, functional programs were excluded if they demonstrated significant inconsistent mediation for any module.

To determine whether functional programs with significant mediating effects act in a sequential manner to influence co-expression module activity, we applied Bayesian network analysis. Bayesian networks were represented as directed acyclic graphs (DAG), in which nodes corresponded to variant carrier status, functional program activity, and co-expression module activity, while edges represented conditional dependencies between variables. Let $X= \left\{ X_{1},X_{2},\ldots,X_{n} \right\}$ denote the set of variables in the network. The join probability distribution was factorized according to the DAG structure as:

$$P\left( X_{1},X_{2},\ldots,X_{n} \right)=\prod_{i=1}^{n} P(X_{i}|Pa\left( X_{i} \right))$$

where $Pa\left( X_{i} \right)$ denotes the parent nodes of variable $X_{i}$. Prior structural restrictions were imposed during network learning: edges directed toward variant carrier status were prohibited, co-expression modules were prohibited from regulating upstream functional programs (informed by AIC values), and direct edges from variant carrier status to co-expression modules were prohibited to enforce candidate mediating paths. Network structures were initially learned using score-based Bayesian network optimization and subsequently refined using automated heuristic structure search. To assess edge stability, bootstrap resampling was performed (*N* = 100 iterations); samples were drawn with replacement, the Bayesian network was relearned under identical structural constraints, and all directed edges were extracted. Edge stability was quantified as:

$$Edge {frequency}_{i\to j}=\frac{N_{i\to j}}{N_{boot}}$$

where $N_{i\to j}$ represents the number of bootstrap iterations in which edge $i\to j$ was observed and $N_{boot}$ represents the total number of bootstrap iterations. Edges detected in at least 50% of bootstrap iterations were retained as candidate pathways linking variants in ALS genes to co-expression module activity.

Finally, we evaluated all candidate pathways linking variants in ALS genes to co-expression module activity using serial mediation analysis to determine whether functional programs exerted significant mediating effects along the inferred paths. Let $X$ denote variant carrier status, $Y$ denote co-expression module activity, and $M_{1},M_{2},\ldots,M_{k}$ denote sequential functional program mediators inferred from the Bayesian network. Serial mediation models were constructed such that each mediator was conditioned on the preceding variable in the inferred path:

$$M_{1}=a_{1}X+C\beta_{1}+ℇ_{1}$$

$$M_{2}=a_{2}M_{1}+C\beta_{2}+ℇ_{2}$$

$$\vdots$$

$$M_{k}=a_{k}M_{k-1}+C\beta_{k}+ℇ_{k}$$

The outcome model was then defined as:

$$Y=bM_{k}+c^{'}X+\sum_{i=1}^{k-1} p_{i}M_{i}+C\gamma+ℇ$$

Where $b$ represents the effect of the terminal mediator on module activity and $p_{i}$ represents parallel contributions from intermediate mediators to the outcome. The serial indirect effect along the inferred pathway was defined as:

${Indirect}_{serial}= a_{1}\times a_{2}\times\cdots\times a_{k}\times b$

Parallel indirect effects were estimated for intermediate mediators directly connected to the outcome:

${Indirect}_{pi}= a_{i}\times p_{i}$

The total indirect effect was defined as:

$$Total indirect effect= {Indirect}_{serial}+ \sum_{i=1}^{k-1} {Indirect}_{pi}$$

And the total effect was defined as:

$$Total effect=c^{'}+Total indirect effect$$

This framework informed whether sequentially connected functional programs significantly mediated the effect of variants in ALS genes on co-expression module activity. Transcriptional pathways exhibiting significant serial effects after Bonferroni correction (*p* < 0.05) were subjected to genetic association analyses. For co-expression modules, only genes with |kME| > 0.6 were queried, thereby excluding genes with weak contributions to module activity.
