## Supplementary File 2 for "Transcriptional pathways of definitive ALS genes implicate novel disease-associated target genes"

**Supplementary Figures 1-10**

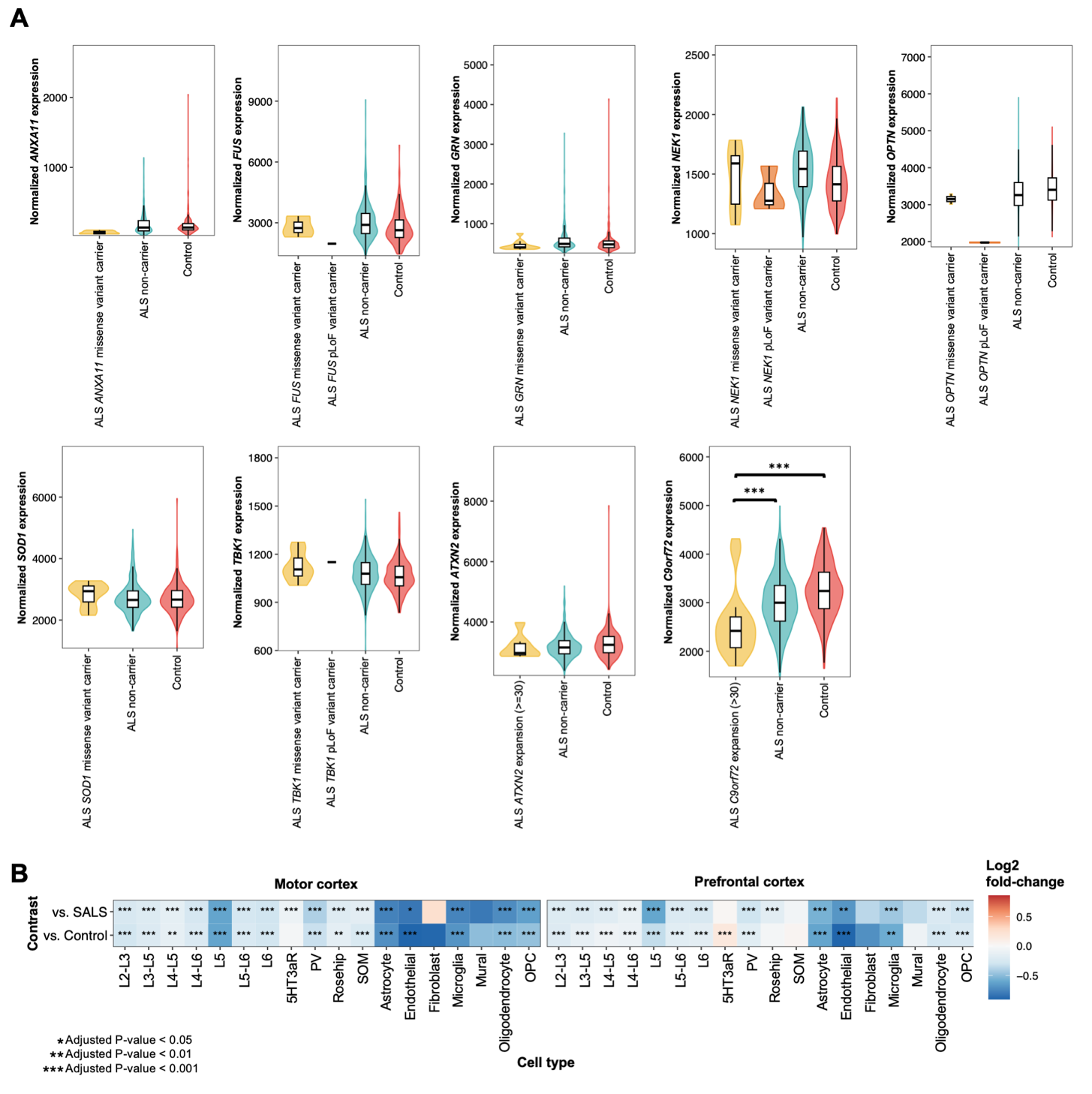

**Supplementary Fig. 1. Expression of definitive ALS genes across variant carrier groups. A)** Expression data are derived from bulk RNA sequencing of induced pluripotent stem cell-derived motor neurons from individuals with ALS and controls. Violin plots show the expression of definitive ALS genes, stratified by variant carrier group. Differential expression between ALS variant carriers versus ALS non-carrier and ALS variant carriers versus controls was assessed using a Wilcoxon rank-sum test. **B)** Expression data are derived from single-nucleus RNA sequencing of the motor and prefrontal cortices of individuals with *C9orf72*-associated ALS (C9ALS), sporadic ALS (SALS), and controls. The heatmap shows the cell type-specific log2 fold-change values in *C9orf72* expression between C9ALS versus SALS and C9ALS versus controls. Differential expression was computed using MAST. P-values presented in panels A and B were adjusted for multiple testing using the Benjamini-Hochberg procedure to control the false discovery rate.

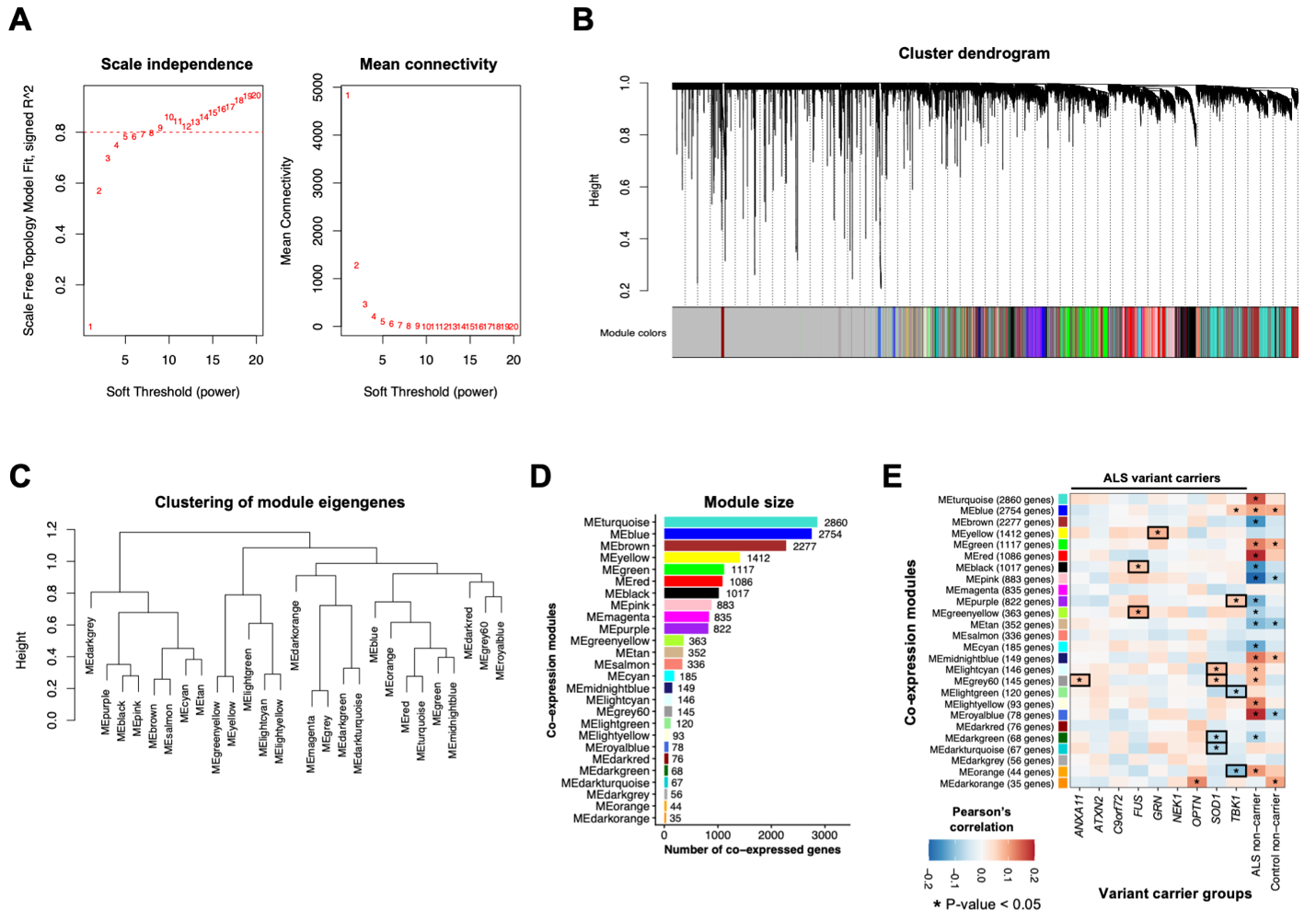

**Supplementary Fig. 2 Overview of the weighted gene co-expression network analysis. A)** Selection of the soft-thresholding power based on scale-free topology fit (left) and mean connectivity (right). A soft-thresholding power of 10 was selected for network construction. **B)** Hierarchical clustering dendrogram of genes, with modules assigned distinct colors. **C)** Clustering dendrogram of module eigengenes based on pairwise similarity of co-expression profiles. **D)** Number of genes assigned to each co-expression module. **E)** Heatmap showing Pearson’s correlation between module eigengenes and variant carrier groups.

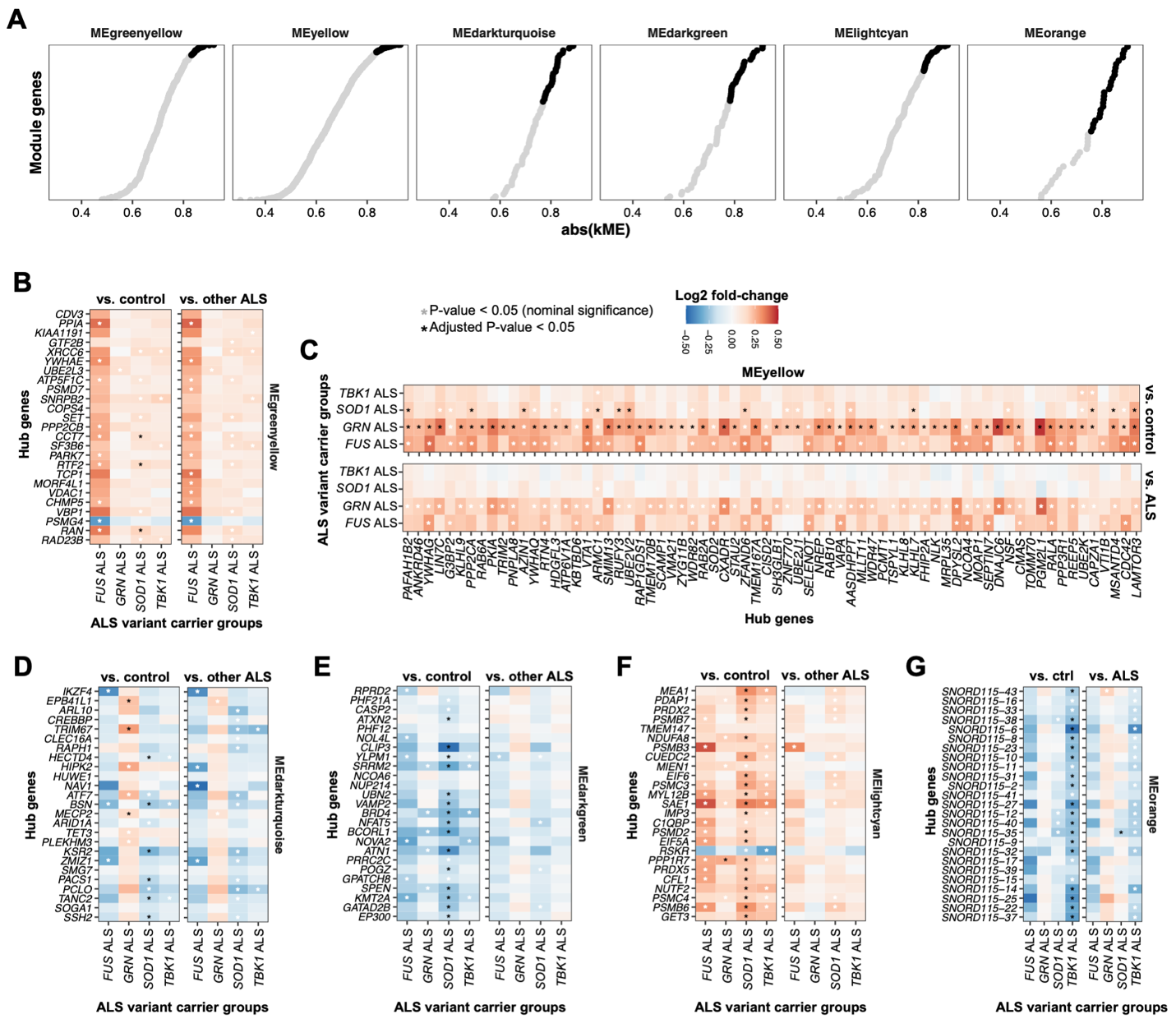

**Supplementary Fig. 3. Characterization of co-expression module hub genes. A)** Distribution of module membership (kME) values across genes within each co-expression module. Genes were ranked by absolute kME, and the top 5% per module (minimum of 25 genes) were selected as hub genes. Heatmaps show log2 fold-change in expression of module hub genes comparing ALS variant carriers versus controls (left) and ALS variant carriers versus ALS non-carriers (right) for **B)** MEgreenyellow, **C)** MEyellow, **D)** MEdarkturquoise, **E)** MEdarkgreen, **F)** MElightcyan, and **G)** MEorange. Differential expression between groups was assessed using a Wilcoxon rank-sum test. P-values were adjusted for multiple testing using the Benjamini-Hochberg procedure to control the false discovery rate.

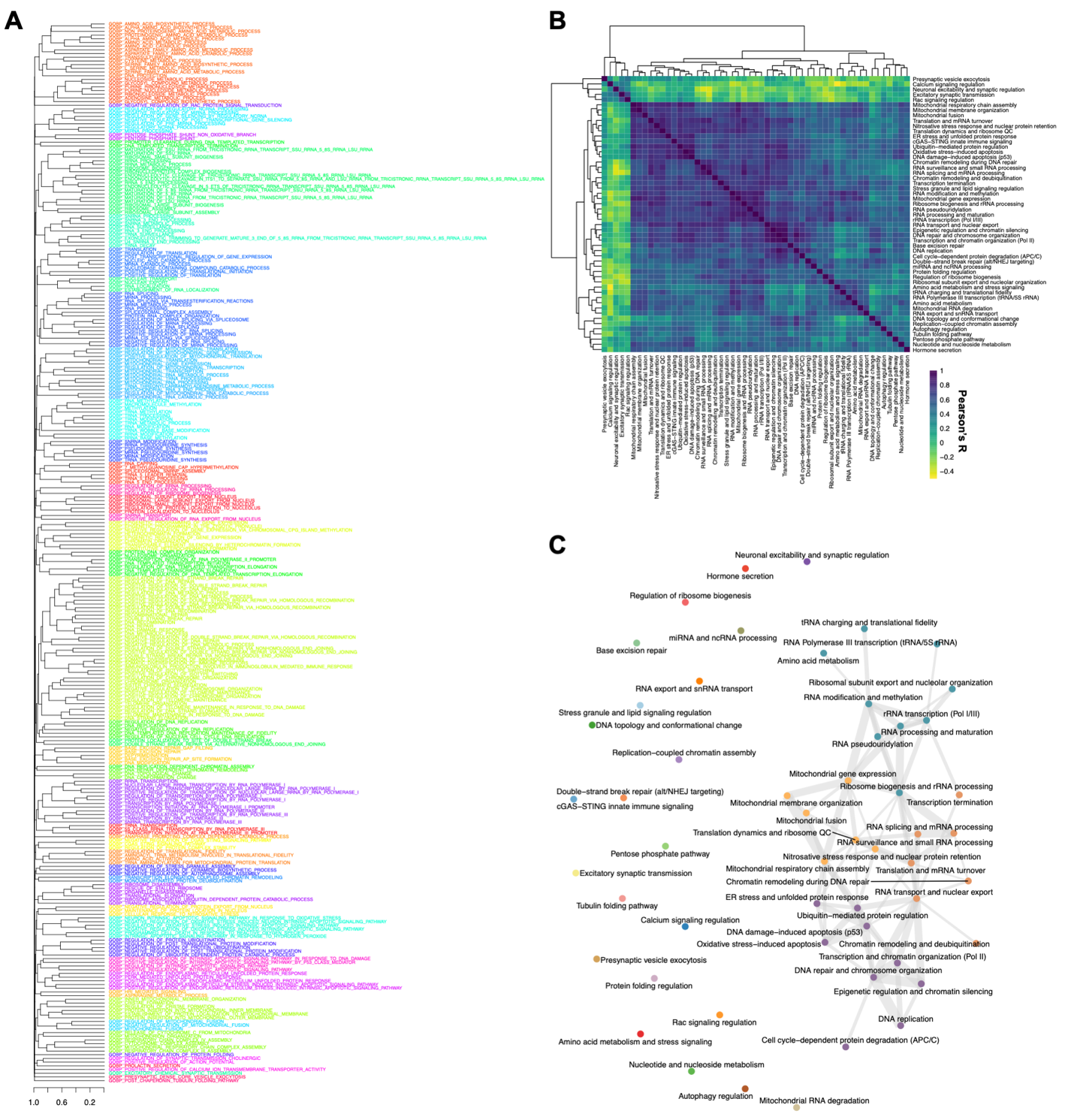

**Supplementary Fig. 4. Construction of functional programs with altered activity in individuals with ALS carrying variants in *SOD1*. A)** Dendrogram showing the hierarchical clustering of Gene Ontology Biological Process (GO:BP) terms based on shared gene membership. **B)** Heatmap showing pairwise Pearson correlations between cluster-level GSVA activity profiles. **C)** Network representation of functional programs derived from hierarchical clustering of the similarity matrix using Euclidean distance and complete linkage. Nodes represent GO:BP clusters, while edges represent correlation strength between clusters.

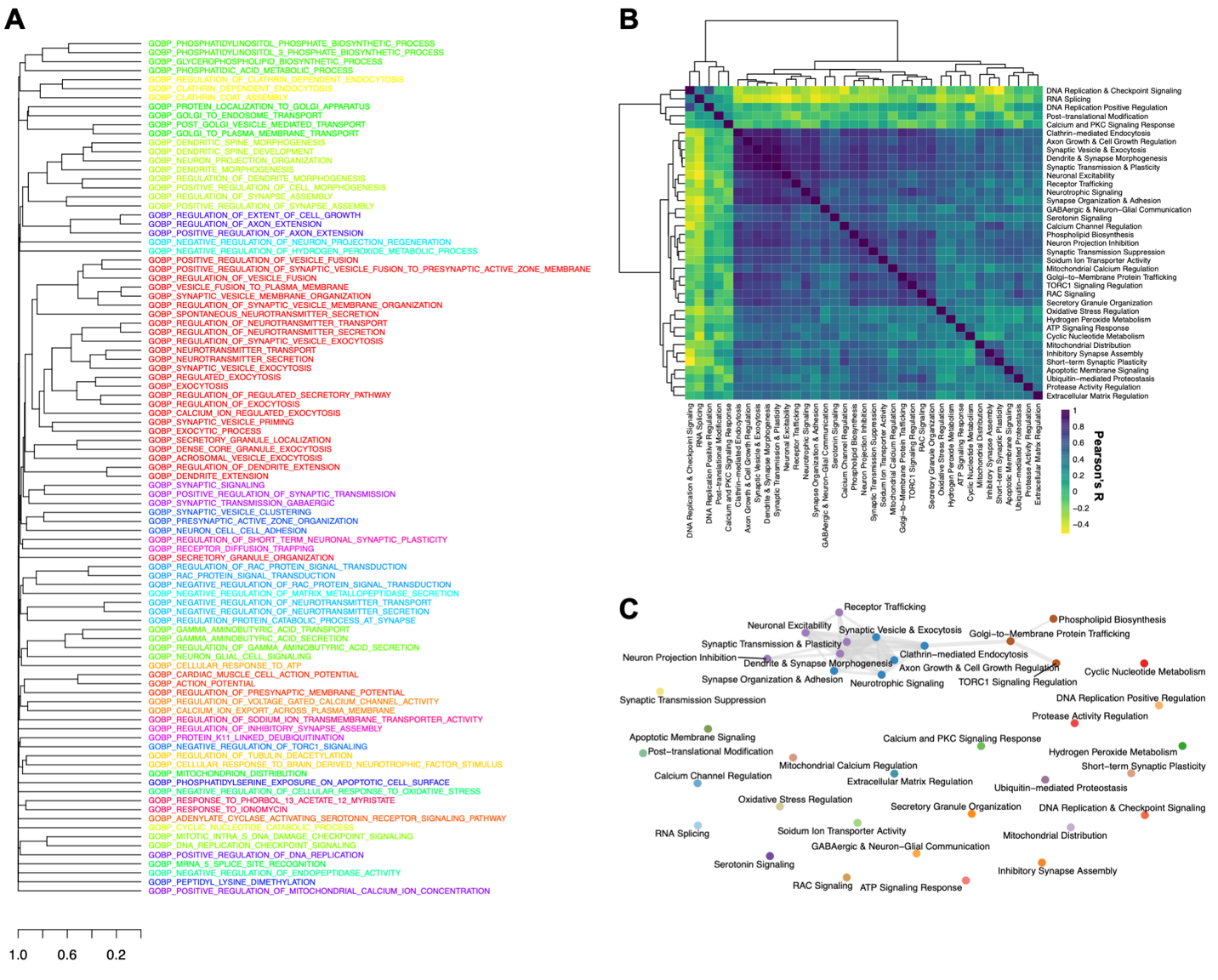

**Supplementary Fig. 5. Construction of functional programs with altered activity in individuals with ALS carrying variants in *GRN*. A)** Dendrogram showing the hierarchical clustering of Gene Ontology Biological Process (GO:BP) terms based on shared gene membership. **B)** Heatmap showing pairwise Pearson correlations between cluster-level GSVA activity profiles. **C)** Network representation of functional programs derived from hierarchical clustering of the similarity matrix using Euclidean distance and complete linkage. Nodes represent GO:BP clusters, while edges represent correlation strength between clusters.

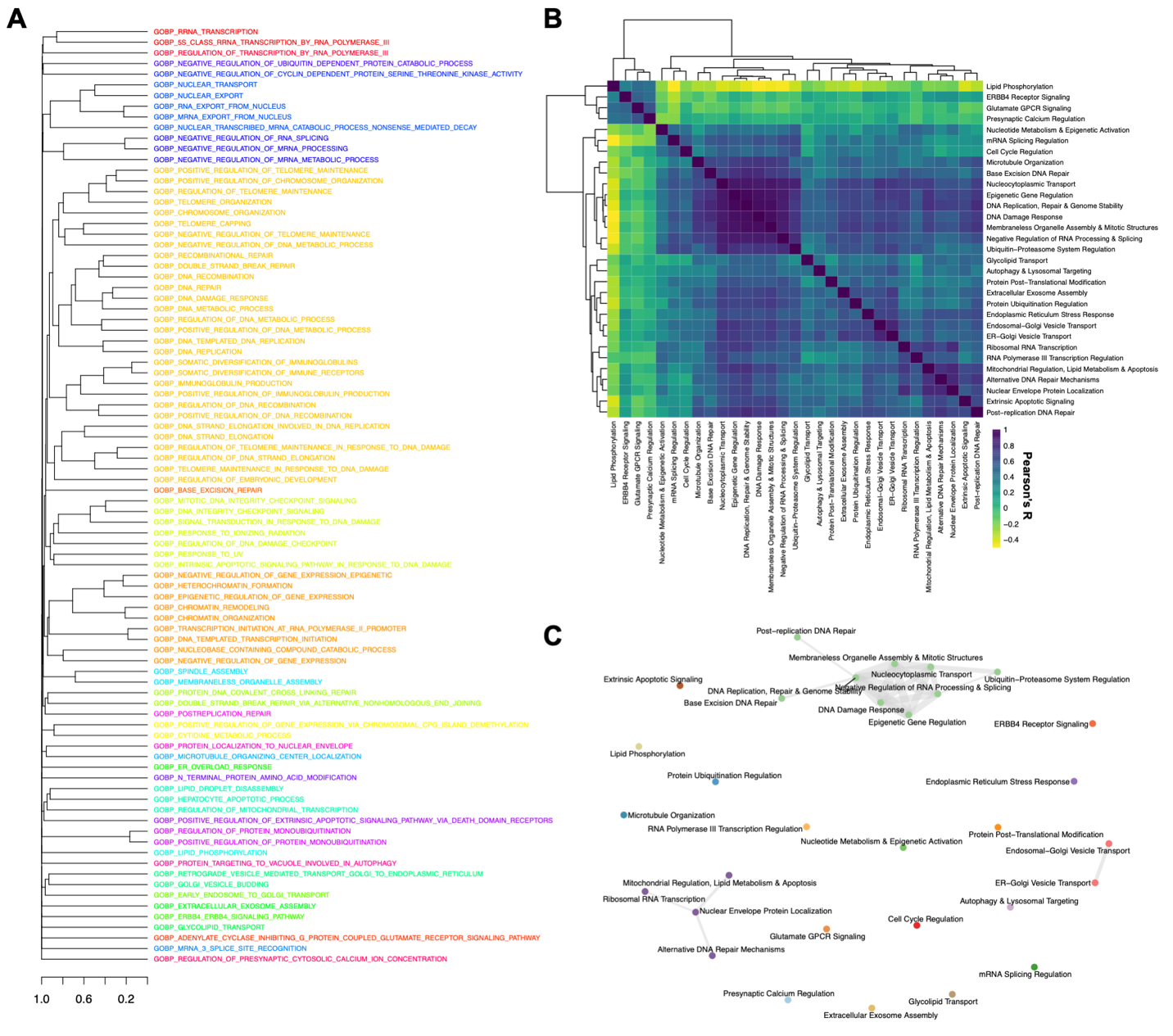

**Supplementary Fig. 6. Construction of functional programs with altered activity in individuals with ALS carrying variants in *TBK1*. A)** Dendrogram showing the hierarchical clustering of Gene Ontology Biological Process (GO:BP) terms based on shared gene membership. **B)** Heatmap showing pairwise Pearson correlations between cluster-level GSVA activity profiles. **C)** Network representation of functional programs derived from hierarchical clustering of the similarity matrix using Euclidean distance and complete linkage. Nodes represent GO:BP clusters, while edges represent correlation strength between clusters.

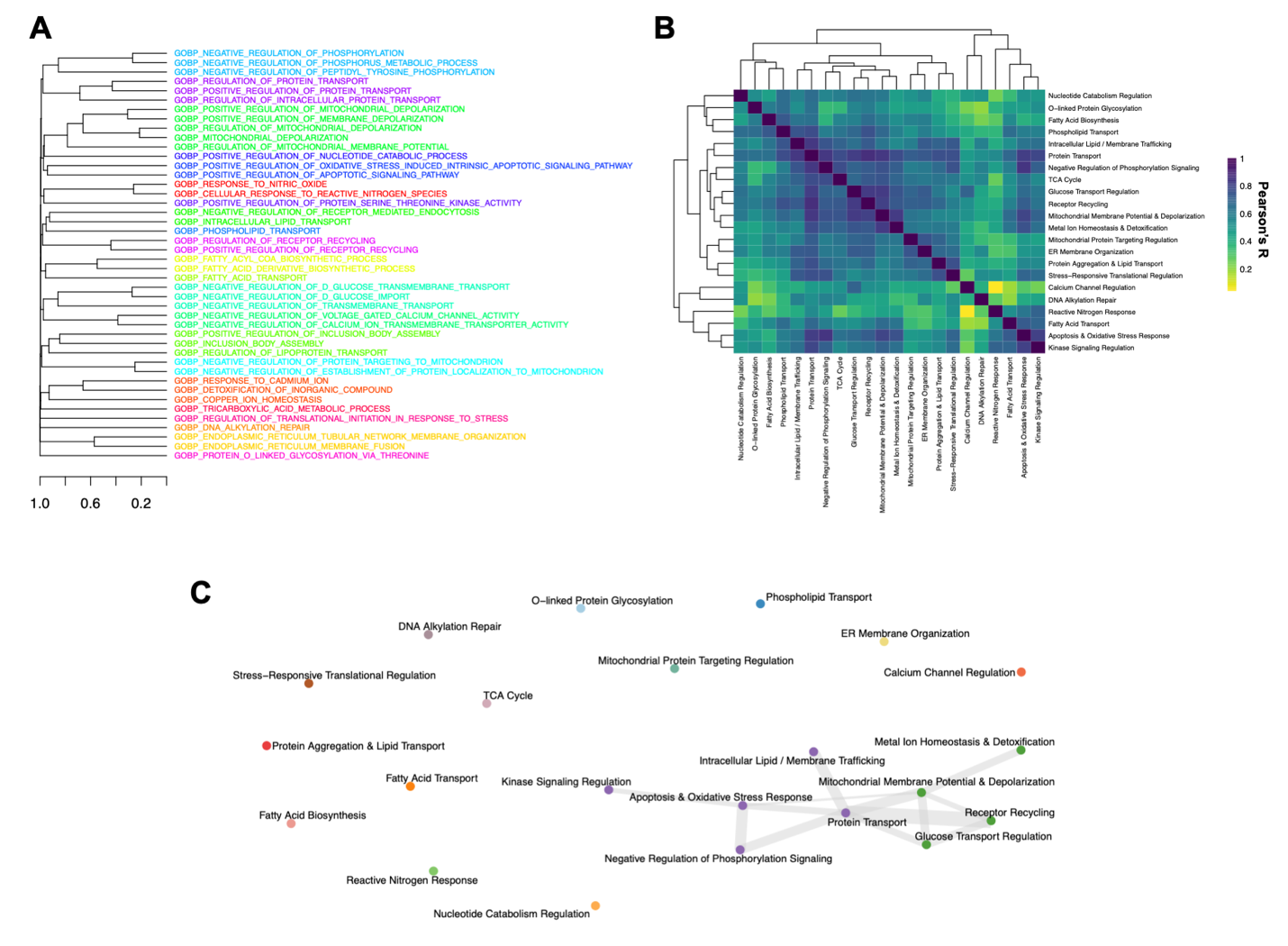

**Supplementary Fig. 7. Construction of functional programs with altered activity in individuals with ALS carrying variants in *FUS*. A)** Dendrogram showing the hierarchical clustering of Gene Ontology Biological Process (GO:BP) terms based on shared gene membership. **B)** Heatmap showing pairwise Pearson correlations between cluster-level GSVA activity profiles. **C)** Network representation of functional programs derived from hierarchical clustering of the similarity matrix using Euclidean distance and complete linkage. Nodes represent GO:BP clusters, while edges represent correlation strength between clusters.

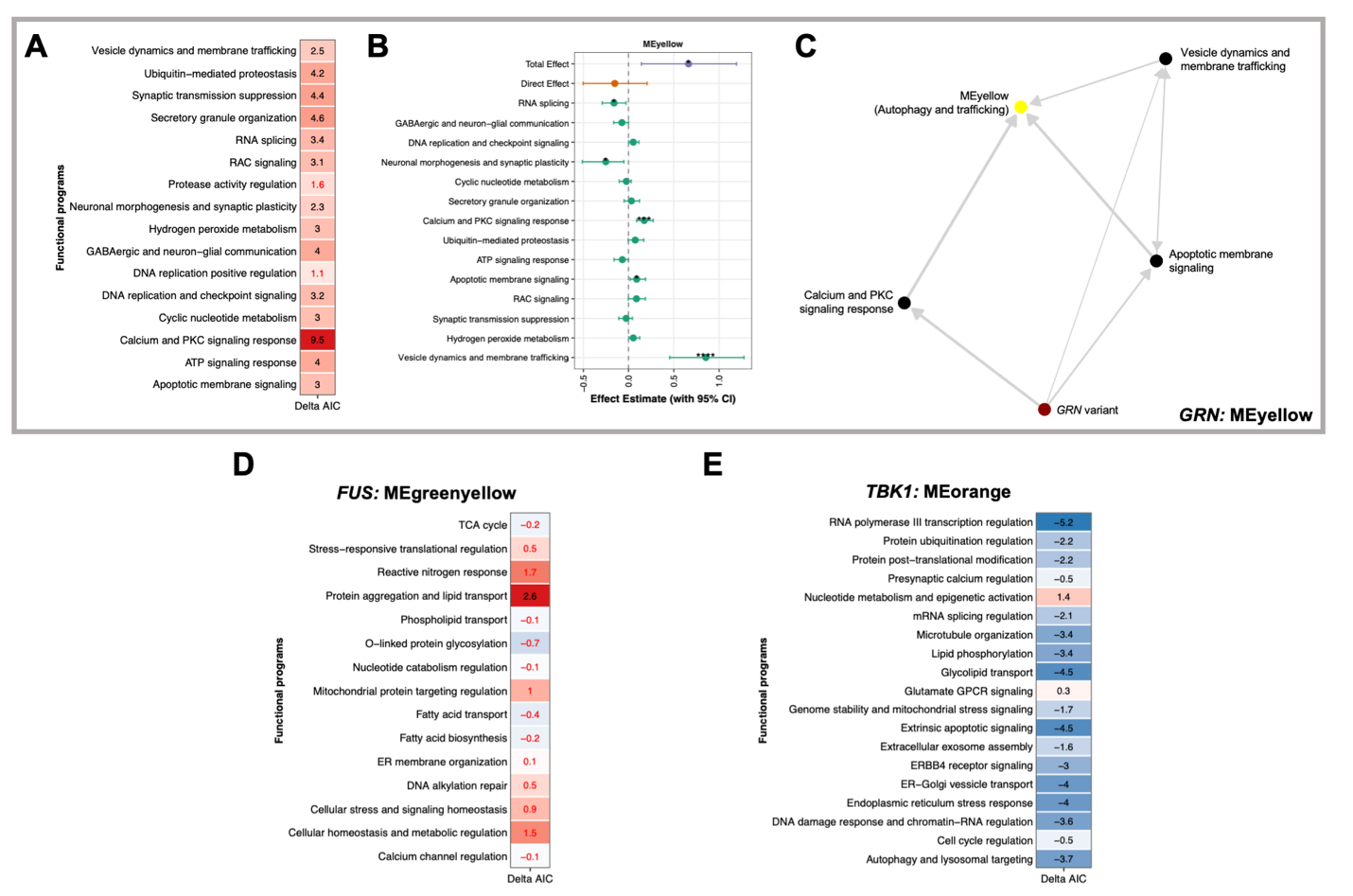

**Supplementary Fig. 8.** Panels A-C correspond to the *GRN*-MEyellow module-trait association. **A)** Heatmap displaying ΔAIC values comparing two competing mediation models in which functional programs were modeled either as mediators of co-expression module activity or as downstream outcomes. Functional programs with ΔAIC > 2, indicating stronger support for the mediator model, were retained for downstream analyses. **B)** Total, direct, and indirect (mediated) effects estimated from parallel mediation models. Functional programs with significant indirect effects concordant with the observed effect of variant carrier status on co-expression module activity were retained for downstream analyses. **C)** Network representation of the Bayesian network analysis used to infer directional relationships among variants in *GRN*, functional programs, and co-expression modules. Nodes represent genetic status, functional programs, or co-expression modules, while edges represent inferred conditional dependency relationships between nodes. Candidate pathways identified from the Bayesian network were subsequently evaluated using serial mediation analysis. **D)** Same as A, but for the *FUS*-MEgreenyellow module trait association.

**E)** Same as A, but for the *TBK1*-MEorange module trait association.

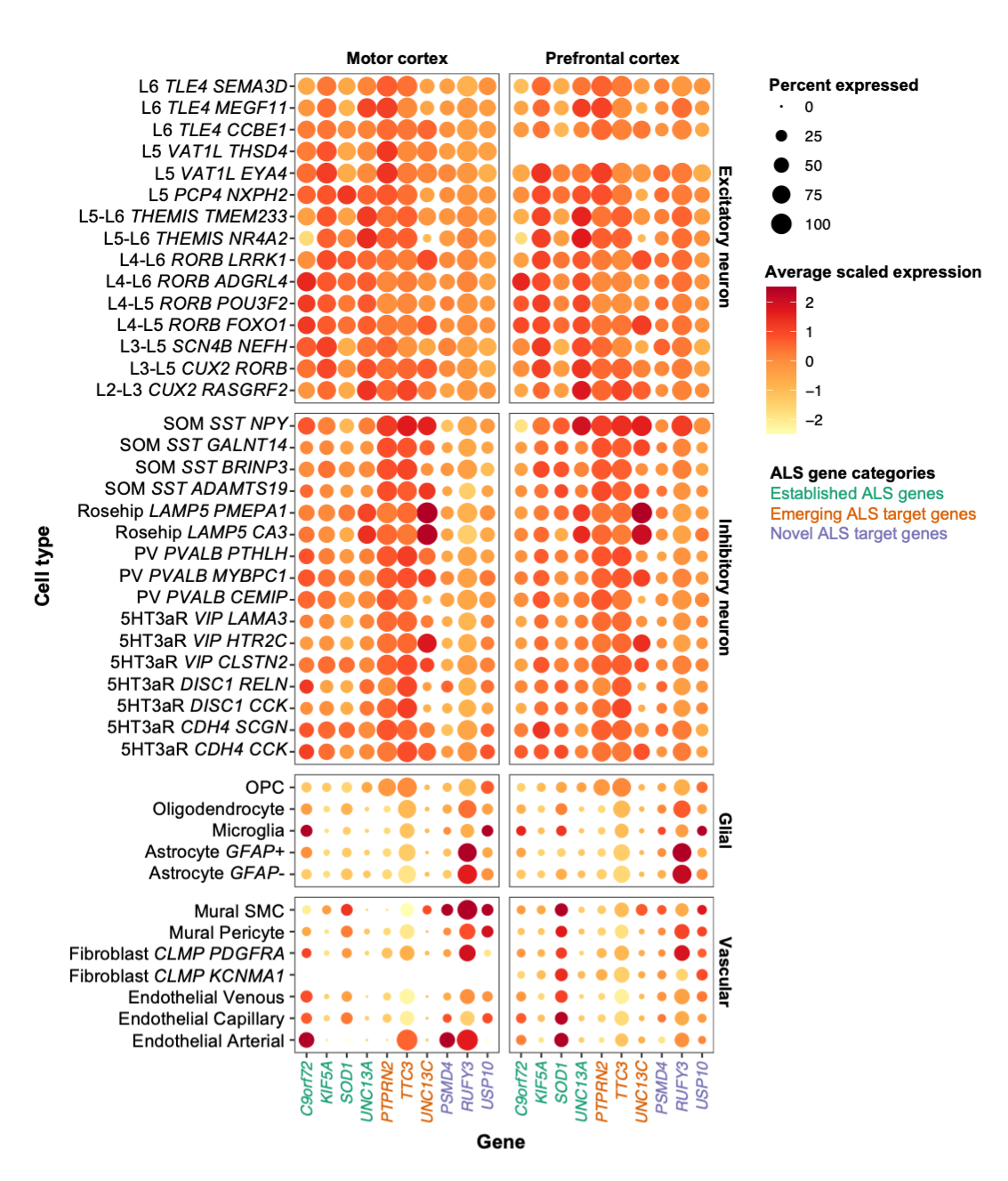

**Supplementary Fig. 9. Expression of ALS target genes across transcriptionally defined cellular subtypes in the motor and prefrontal cortices.** The single-nucleus RNA sequencing data are derived from the motor and prefrontal cortices (MCx/FCx) of neurologically healthy controls. The dot plot shows the average scaled expression and percent expression of established ALS genes (green), emerging ALS target genes (orange), and novel ALS target genes (purple) across cellular subtypes comprising the MCx and FCx.

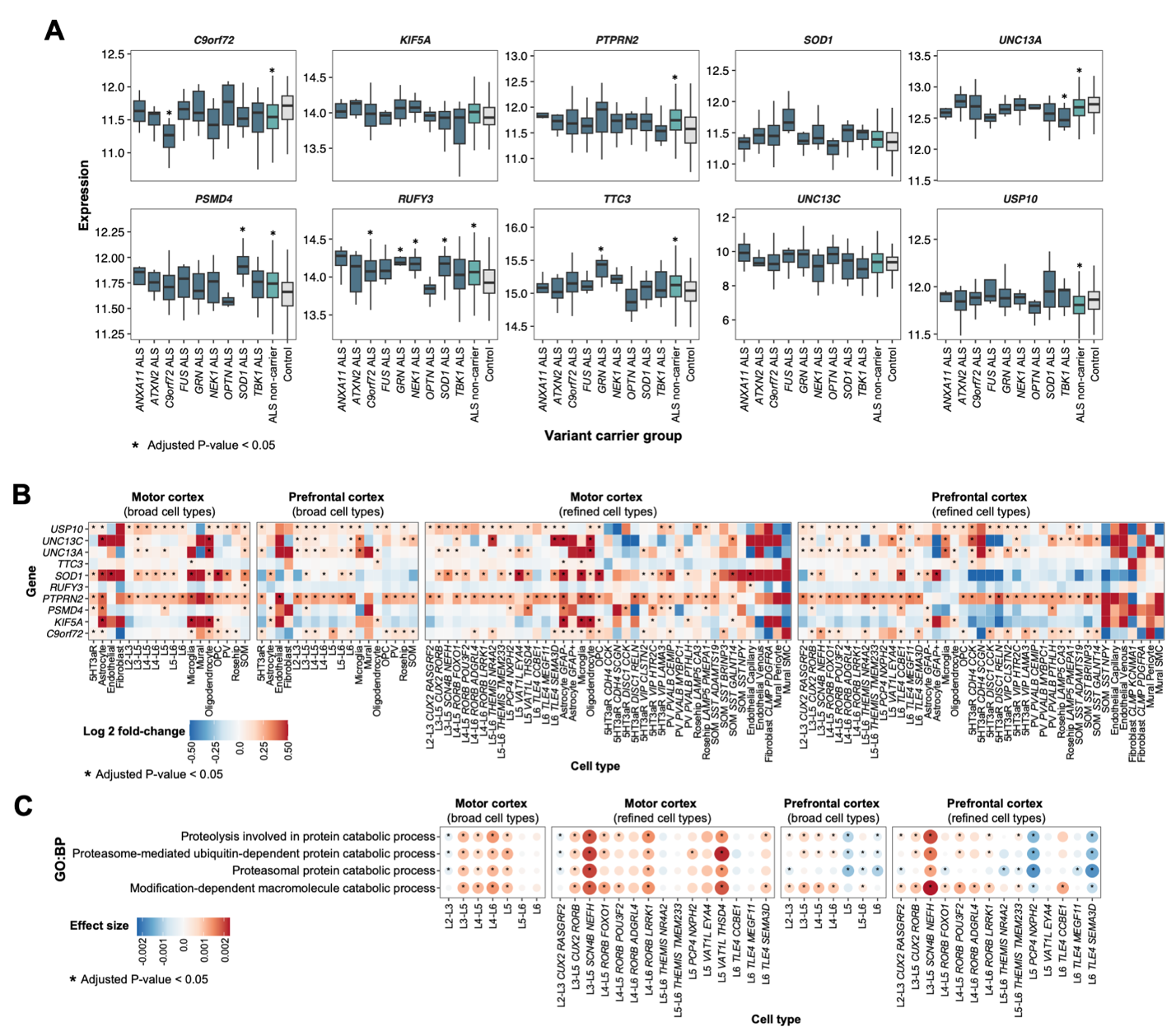

**Supplementary Fig. 10. Gene expression of ALS target genes at a bulk and single-nuclei resolution. A)** Target gene expression across variant carrier groups in bulk RNAseq data from induced pluripotent stem cell-derived motor neurons. Differential expression between ALS variant carriers versus controls was assessed using a Wilcoxon rank-sum test. **B-C**) The single-nucleus RNA sequencing data are derived from the motor and prefrontal cortices (MCx/FCx) of individuals with sporadic ALS (SALS) and neurologically healthy controls. **B)** Differential expression of target genes across cellular populations between SALS and controls, stratified by brain region and cell type resolution. Differential expression was computed using MAST. **C)** Dot plot showing differential activity of Gene Ontology Biological Process (GO:BP) pathways containing *PSMD4* across cellular populations quantified using *UCell*, stratified by brain region and cell type resolution. Differential pathway activity between SALS and controls was assessed using multivariable linear regression. P-values presented in panels A-C were adjusted for multiple testing using the Benjamini-Hochberg procedure to control the false discovery rate.
